# Measuring the Unmeasurable: A Diagnostic Sensor for AI Reasoning Pathology in Sequential Clinical Decision-Making

**DOI:** 10.64898/2026.03.27.26349583

**Authors:** Stella X. Wang

## Abstract

Large Language Models achieve impressive accuracy on medical benchmarks that present clinical information as complete vignettes, but their behavior under sequential information delivery, the standard mode of real clinical practice, is poorly characterized. We conduct a three-condition ablation study (N=50 clinical cases from published case reports in PubMed Central, 150 total runs) using claude-sonnet-4-20250514 to investigate what happens when diagnostic information arrives in stages rather than all at once. We introduce a novel 5+2 scoring rubric measuring seven dimensions of reasoning quality beyond binary accuracy, and a 6-code failure mode taxonomy enabling mechanistic root-cause analysis of diagnostic failures.

We document **Convergence Regression (CR)**: a systematic failure mode where models correctly identify diagnoses at intermediate reasoning stages but abandon them when subsequent evidence triggers pattern-matching to alternative diagnoses. Under unstructured sequential delivery, models access the correct diagnosis in 90% of cases but retain it in only 60%, creating a 30% **Access-Stability Dissociation** invisible under single-shot evaluation. A structured scaffold, the Sequential Information Prioritization Scaffold (SIPS), eliminates this gap entirely through forced hypothesis accountability: 80% access, 80% final accuracy, 0% Convergence Regression. We term this the **SIPS Retention Effect**. However, scaffolding reduces top-1 accuracy from 60% to 40%, a **Convergence Hesitancy Paradox** establishing that retention and convergence are architecturally distinct reasoning tasks requiring separate mechanisms.

We propose that structured scaffolding functions as a **diagnostic sensor** for reasoning pathology rather than an accuracy intervention: it makes failure modes visible, classifiable, and auditable. We demonstrate that our measurement instruments operationalize WHO and FDA governance requirements for AI transparency, accountability, and safety into quantifiable scores. We release the complete framework, including the 5+2 rubric, 6-code taxonomy, scaffold specification, and 210-score matrix with adjudication rationale, as a reusable audit instrument for evaluating LLM reasoning behavior in any sequential reasoning context.

## 1. Introduction

### 1.1 The Clinical Reasoning Gap

Recent advances in Large Language Models have produced remarkable performance on standardized medical benchmarks. GPT-4 achieved approximately 86% on the United States Medical Licensing Examination (Nori et al., 2023). Med-PaLM 2 reached 86.5% on MedQA and demonstrated strong performance across the MultiMedQA benchmark suite (Singhal et al., 2023). Med-Gemini pushed the state of the art to 91.1% on MedQA (Saab et al., 2024), and subsequent models have continued this trajectory, with GPT-5 reporting 95.84% on MedQA in 2025. These results have generated significant interest in deploying LLMs as clinical decision-support tools.

However, a fundamental disconnect exists between these benchmark evaluations and the reality of clinical practice. Medical benchmarks, including MedQA, USMLE-style examinations, and clinical vignette assessments, **present information as complete, static vignettes**. The model receives a fully specified clinical scenario and produces a diagnosis from the totality of available evidence. This is not how clinical reasoning works. In real-world medicine, diagnostic information arrives in **sequential stages**: an initial presentation in the emergency department, followed by a physical examination, then incremental laboratory results, imaging findings, and specialty consultations. At each stage, the clinician must update, refine, or abandon existing hypotheses based on new evidence that may support, contradict, or redirect the diagnostic trajectory.

This temporal dimension of clinical reasoning introduces cognitive demands that static benchmarks do not measure. Human clinicians are known to exhibit systematic reasoning failures under sequential information delivery, including anchoring bias (Croskerry, 2003), premature closure (Graber et al., 2005), and confirmation bias, with diagnostic errors contributing to an estimated 800,000 deaths or permanent disabilities annually in the United States alone (Topol, 2024). Whether LLMs exhibit analogous reasoning pathologies under sequential delivery, and whether these pathologies can be measured and mitigated, remains poorly characterized.

### 1.2 The Problem We Address

This paper asks two questions. First: **what happens to LLM diagnostic reasoning when information is delivered sequentially, as in real clinical practice?** Existing evaluations cannot answer this question because they present complete information in a single prompt. By delivering the same clinical cases in four sequential stages, each potentially introducing evidence that supports, contradicts, or redirects the current differential, we expose the model to the temporal uncertainty that defines real diagnostic workflows.

Second: **can structured scaffolding prevent the reasoning failures that emerge under sequential delivery?** If sequential delivery introduces systematic instability, the follow-up question is whether structural interventions can restore stability without sacrificing the broader exploratory benefits that sequential processing provides.

We approach these questions through a **measurement thesis**: we propose that scaffolding should be understood not primarily as a treatment for poor reasoning, but as a **measurement instrument** that makes reasoning pathology visible and quantifiable. Without structural accountability, failure modes operate silently: a model may find the correct diagnosis at an intermediate stage and then lose it, with no trace of the regression in the final output. With structural accountability, these failures become auditable events. The scaffold does not merely improve performance; it reveals the cognitive architecture of the model’s reasoning process.

### 1.3 Approach and Preview of Findings

We conduct a three-condition ablation study on N=50 clinical cases derived from published case reports indexed in PubMed Central (PMC), using a single model (claude-sonnet-4-20250514) under deterministic settings. The three conditions isolate two variables: information delivery mode (single-shot vs. sequential) and structural scaffolding (absent vs. present):

- **C1 (Single-Shot Baseline):** The complete clinical vignette is delivered in a single prompt. The model produces one diagnostic output from the totality of available evidence.
- **C2 (Sequential, No Scaffold):** The same clinical information is delivered across four sequential stages. The model updates its differential at each stage without structural requirements beyond ’update your diagnosis.’
- **C3 (Sequential, SIPS-Scaffolded):** The same four-stage delivery, but with the Sequential Information Prioritization Scaffold (SIPS): a structured prompting framework that enforces forced differential ranking, explicit hypothesis rotation documentation, and convergence status tracking at every stage.

All 30 traces from a 10-case deep-analysis subset were scored using a novel 5+2 rubric measuring seven dimensions of reasoning quality beyond binary accuracy, and all incorrect outcomes were classified using a 6-code failure mode taxonomy identifying the mechanism of each diagnostic failure.

Our findings reveal a phenomenon we term **Convergence Regression**: under sequential delivery without scaffolding, the model accesses the correct diagnosis at intermediate stages at a substantially higher rate than it retains the correct diagnosis in its final output. This **Access-Stability Dissociation**, invisible under single-shot evaluation, represents a qualitatively new category of LLM reasoning failure. We further demonstrate that structured scaffolding eliminates this gap entirely through what we term the **SIPS Retention Effect**: forced hypothesis accountability prevents the silent abandonment of correct diagnoses. However, this retention comes at the cost of reduced top-1 convergence, a trade-off we term the **Convergence Hesitancy Paradox**, revealing that retention and convergence are architecturally distinct reasoning tasks requiring different interventions.

### 1.4 Contributions

This paper makes the following contributions:

- **We document Convergence Regression (CR):** a novel failure mode where LLMs find correct diagnoses at intermediate stages of sequential reasoning but systematically abandon them when subsequent information triggers pattern-matching to alternative diagnoses. CR represents a failure of reasoning stability, not reasoning capability, and is invisible under standard single-shot evaluation.
- **We introduce the 5+2 scoring rubric:** a multi-dimensional measurement framework that assesses reasoning quality beyond binary accuracy. Five core dimensions (Diagnostic Accuracy, Reasoning Depth, Calibration, Hypothesis Tracking, Step Adherence) and two diagnostic dimensions (Early Convergence, Anchoring Resistance) provide a seven-axis profile of reasoning behavior applicable to any LLM.
- **We demonstrate the SIPS Retention Effect:** structured scaffolding with forced hypothesis accountability eliminates the Convergence Regression gap through three structural barriers (Visibility, Justification, Convergence Tracking). The scaffold converts an unstable reasoner into a stable one at the cost of reduced top-1 convergence, establishing the architectural boundary between retention and convergence mechanisms.
- **We provide a 6-code failure mode taxonomy:** a mechanistic classification system (KV, RF, SD, PC, LM, CR) that enables root-cause analysis of LLM diagnostic failures. Each code identifies a distinct failure type with different architectural remediation requirements, enabling targeted intervention rather than generic accuracy optimization.
- **We release our complete measurement framework as a reusable audit instrument:** the 5+2 rubric, 6-code taxonomy, SIPS scaffold specification, and full 210-score matrix with adjudication rationale. We demonstrate that these instruments operationalize abstract AI governance principles (WHO Transparency, Accountability, Safety requirements) into quantifiable, reproducible measurements, establishing a path from technical evaluation to regulatory compliance.

The remainder of this paper is organized as follows. Section 2 reviews related work across four domains (LLM medical diagnosis, cognitive biases in clinical reasoning, structured prompting, and test-time compute scaling) and establishes the clinical safety and governance context. Section 3 details the experimental methods including dataset construction, ablation design, SIPS architecture, and the 5+2 rubric specification. Section 4 presents results across eight subsections. Section 5 discusses implications, the retention-convergence trade-off, limitations, and future work. Section 6 concludes. Section 7 addresses broader impact and the measurement-as-moat thesis.

## 2. Related Work

We situate this work at the intersection of four research streams: LLM performance on medical benchmarks, cognitive biases in clinical reasoning, structured prompting architectures, and test-time compute scaling. Each stream contributes a critical dimension to our study, and each reveals a gap that our work addresses.

### 2.1 LLMs in Medical Diagnosis

The evaluation of Large Language Models in medicine has relied heavily on static, single-shot benchmarks. Frontier models have demonstrated exceptional performance on these assessments: GPT-4 achieved approximately 86% on USMLE-style questions (Nori et al., 2023), Med-PaLM 2 reached 86.5% on MedQA and demonstrated strong performance across the MultiMedQA benchmark suite (Singhal et al., 2023), and Med-Gemini pushed the state of the art to 91.1% on MedQA (Saab et al., 2024). Recent iterations have continued this trajectory, with GPT-5 reportedly achieving 95.84% accuracy on MedQA in 2025. These results have generated considerable enthusiasm about the clinical potential of LLMs.

However, as noted in the NEJM AI (2025) clinical reasoning assessment framework, these benchmarks present clinical information as **completed vignettes** rather than the sequential, evolving data streams encountered in actual clinical practice. A model processing a complete vignette performs a fundamentally different cognitive task than a model updating its differential as new evidence arrives across temporal stages. The former requires pattern-matching to a fixed information set; the latter requires hypothesis management under uncertainty, with each new data point potentially confirming, contradicting, or redirecting the diagnostic trajectory.

This distinction is not merely theoretical. Our results demonstrate that models achieving equivalent accuracy under single-shot evaluation exhibit dramatically different reasoning dynamics under sequential delivery, including failure modes that are invisible under static evaluation. The Access-Stability Dissociation documented in this paper is a direct consequence of the gap between how models are evaluated (complete vignettes) and how they will be deployed (sequential information delivery).

### 2.2 Cognitive Biases in Clinical Reasoning

Diagnostic error research has long identified specific cognitive biases that lead to clinical failure. Croskerry (2003) established the foundational importance of cognitive errors in diagnosis and clinical decision-making. Graber et al. (2005) identified **premature closure**, the tendency to narrow a differential too early, as the most prevalent cognitive error in internal medicine, present in 74% of diagnostic failures. Berner and Graber (2008) documented the persistence of overconfidence as a contributing factor to diagnostic error, a finding with direct relevance to our Calibration dimension.

These failures are commonly analyzed through Kahneman’s (2011) dual-process theory, which distinguishes between fast, intuitive pattern-matching (System 1) and slow, analytical deliberation (System 2). Diagnostic errors disproportionately arise when System 1 processing produces a premature hypothesis that System 2 fails to override. Recent work has extended this framework to LLMs: a 2025 study in *Nature npj Digital Medicine* documented that clinical LLMs exhibit anchoring biases consistent with autoregressive processing, and a 2024 JAMA Network Open randomized controlled trial measured the influence of LLM outputs on physician diagnostic reasoning, finding significant automation bias effects.

We extend this literature by documenting **Convergence Regression (CR)**, a failure mode that is related to but distinct from the established cognitive biases. CR is not anchoring (the model does move away from its initial hypothesis), not premature closure (the model does consider the correct diagnosis), and not confirmation bias (the model does process contradicting evidence). CR is a failure of *sequential stability*: the model reaches the correct diagnosis at an intermediate stage and then actively abandons it when subsequent evidence triggers pattern-matching to a more textbook-sounding alternative. This represents a novel contribution to the diagnostic error literature, establishing that LLMs exhibit cognitive pathologies that parallel but do not replicate those documented in human clinicians.

### 2.3 Structured Prompting and Chain-of-Thought

To improve LLM reasoning, researchers have developed various prompting architectures that elicit structured thought processes. Chain-of-Thought (CoT) prompting (Wei et al., 2022) enables models to decompose complex problems into intermediate reasoning steps, substantially improving performance on arithmetic, commonsense, and symbolic reasoning tasks. Tree of Thoughts (Yao et al., 2023) extends CoT to explore multiple reasoning branches, allowing the model to consider alternative paths before committing to a conclusion. ReAct (Yao et al., 2022) further synergizes reasoning with external actions, enabling models to interleave thought and environmental interaction. More recently, Med-MCTS (2024) has treated medical diagnosis as a sequential decision-making problem amenable to Monte Carlo Tree Search, framing diagnostic reasoning as a planning problem with explicit state transitions.

SIPS diverges from these approaches in a fundamental way. CoT, Tree of Thoughts, and ReAct are **generation mechanisms**: they elicit reasoning steps that the model might not otherwise produce. SIPS is a **retention mechanism**: it does not elicit new reasoning steps but forces explicit accountability for hypothesis changes across sequential stages. The distinction matters because the failure mode we address, Convergence Regression, is not a failure of generation (the model successfully generates the correct hypothesis) but a failure of retention (the model abandons it). Standard prompting improvements that enhance generation quality do not address retention failures because the correct hypothesis is already being generated; the problem is that it is being lost.

SIPS’s forced differential documentation, rotation justification requirements, and convergence status tracking create structural barriers against silent hypothesis abandonment. This positions SIPS not as a competitor to CoT or Tree of Thoughts but as a complementary architecture addressing an orthogonal failure mode. A system could combine CoT (for generation quality) with SIPS (for retention stability) to address both failure categories simultaneously.

### 2.4 Test-Time Compute Scaling

The emergence of inference-time scaling has established that allocating additional compute during the reasoning phase can improve performance on complex tasks. Snell et al. (2024) demonstrated that scaling test-time compute yields positive returns for reasoning-heavy problems, and Microsoft’s (2025) work on inference-time scaling has further explored this approach for complex task solving. These findings have generated interest in ’thinking longer’ as a general strategy for improving LLM reasoning quality.

Our work identifies a critical distinction within this literature: the difference between **unstructured overthinking** and **structured deliberation**. In our experiments, unstructured sequential delivery (C2) consumed 3.1 times the tokens of single-shot delivery (C1) with zero improvement in final accuracy. The model thought longer but not better. The additional tokens went toward exploring hypotheses (90% access rate) that were subsequently abandoned (60% final accuracy). By contrast, the SIPS scaffold (C3) consumed a more modest 1.28 times the tokens of C2, yet achieved a 20 percentage-point improvement in final top-3 accuracy.

This result suggests that the return on inference-time tokens is not determined by their *volume* but by their *structure*. Unstructured scaling produces diminishing returns: more tokens, more exploration, but no improvement in outcome stability. Structured scaling produces positive returns: the scaffold’s overhead (forced documentation, convergence tracking, rotation justification) converts tokens from wasted exploration into productive accountability. This finding has implications beyond our specific study: any application of test-time compute scaling should consider whether the additional compute is structurally directed toward the specific reasoning failure being addressed, rather than simply allocated for extended generation.

### 2.5 Clinical Safety and AI Governance

The preceding sections document what LLMs can do in clinical reasoning tasks. A separate question, increasingly urgent, is what LLMs *should be required to demonstrate* before deployment in clinical workflows. Emerging governance frameworks from the World Health Organization, the U.S. Food and Drug Administration, and clinical safety researchers establish requirements that current evaluation practices fail to meet. Our work addresses this gap directly: the 5+2 rubric and 6-code taxonomy are, to our knowledge, the first instruments that operationalize abstract governance principles into quantifiable, reusable measurements of LLM reasoning behavior.

#### 2.5.1 The Governance Landscape: Requirements Without Instruments

The WHO’s 2021 guidance on ethics and governance of AI for health established six core principles, three of which bear directly on LLM clinical reasoning: **transparency and explainability** (Principle 3: the internal logic of AI systems must be accessible to clinicians and regulators), **accountability** (Principle 4: organizations must be responsible for the outputs and consequences of their AI systems), and **safety** (Principle 2: AI must promote human well-being and not introduce unmonitored risks) (WHO, 2021). The WHO’s 2024 update on large multi-modal models reinforced these principles with over 40 specific recommendations for governments, technology companies, and healthcare providers, explicitly addressing generative AI in clinical decision-support contexts (WHO, 2024).

The FDA’s evolving regulatory framework for AI/ML-enabled clinical decision support similarly emphasizes audit trails, traceability, and the capacity to document algorithmic reasoning. The 2024 final guidance on predetermined change control plans and the 2025 draft guidance on AI-enabled device lifecycle management require immutable audit trails and validation strategies that capture not just what a model outputs, but *how* it arrived at that output (FDA, 2024; FDA, 2025).

Despite the clarity of these requirements, **no standardized instruments exist to fulfill them for LLM clinical reasoning.** Current evaluation practices rely almost exclusively on accuracy benchmarks: MedQA scores, USMLE pass rates, diagnostic concordance. These metrics answer whether the model produced the right answer. They do not answer *how* the model reasoned, *whether* it considered alternatives, *when* it changed its mind, or *why*. This is the observability gap: governance frameworks demand reasoning transparency, but the field lacks measurement tools to provide it.

#### 2.5.2 The Automation Bias Problem: Why Observability Matters

Recent empirical work has sharpened the urgency of this gap. Topol (2024) documented that nearly 800,000 Americans die or are permanently disabled annually from diagnostic errors, framing AI-assisted diagnosis as a potential path toward eradication of these failures. However, subsequent research on automation bias reveals a critical dependency: the safety of AI-assisted diagnosis hinges on the *quality of the reasoning trace*, not merely the accuracy of the final output.

A 2025 study published through medRxiv demonstrated that erroneous LLM recommendations significantly degraded physicians’ diagnostic performance through automation bias, even among AI-trained physicians. Physicians receiving error-free AI recommendations achieved 84.9% diagnostic accuracy, but those exposed to flawed recommendations scored only 73.3%, indicating that clinicians defer to LLM outputs without independently auditing the reasoning process. Critically, providing model explainability did not mitigate this effect: the mere presence of a confident-sounding rationale induced trust regardless of correctness.

This finding has direct implications for our work. Convergence Regression, the failure mode we document in Section 4.3, is particularly dangerous in an automation bias context. When a model initially identifies the correct diagnosis (reaching 90% access under C2) and then abandons it in favor of a more textbook-sounding alternative, **the final output is a confident, well-reasoned wrong answer.** A clinician experiencing automation bias would see a plausible rationale and accept the incorrect conclusion. Without an observability layer that flags the regression, the silent abandonment of the correct diagnosis becomes invisible to human oversight.

#### 2.5.3 Operationalizing Governance: The 5+2 Rubric as Audit Instrument

We propose that the 5+2 rubric introduced in this paper represents a concrete operationalization of the abstract governance requirements outlined above. The following mapping connects each WHO governance principle to specific rubric dimensions:

**Table 1.** Mapping of WHO AI governance principles to specific 5+2 rubric dimensions and the 6-code failure taxonomy. Each governance requirement is operationalized through a concrete, quantifiable measurement.

| WHO Principle | Governance Requirement | 5+2 Rubric Dimension | What It Measures |
| --- | --- | --- | --- |
| **Transparency (P3)** | Internal logic accessible to clinicians | **Hypothesis Tracking (1-5)** | Whether the model documents its differential evolution: additions, removals, promotions, demotions, and explicit rationale for each change. |
| **Transparency (P3)** | Reasoning process intelligible to regulators | **Reasoning Depth (1-5)** | Whether the model connects evidence to pathophysiological mechanisms rather than pattern-matching to surface features. |
| **Accountability (P4)** | Outputs traceable and auditable | **Step Adherence (1-5)** | Whether the model follows the scaffold's structural requirements, producing auditable traces at every stage. Without structural compliance, audit is impossible. |
| **Accountability (P4)** | Immutable audit trail of decisions | **Convergence Regression (CR code)** | Whether previously confirmed diagnoses are silently abandoned. CR detection is only possible when the audit trail forces explicit documentation of hypothesis changes. |
| **Safety (P2)** | No unmonitored reasoning risks | **Calibration (1-5)** | Whether the model's expressed confidence tracks the strength of available evidence. Miscalibrated confidence magnifies automation bias risk. |
| **Safety (P2)** | Bias detection and mitigation | **Anchoring Resistance (1-5)** | Whether the model updates its reasoning when new evidence contradicts initial hypotheses. Low anchoring resistance indicates systematic cognitive bias. |
| **Safety (P2)** | Identify failure modes pre-deployment | **6-Code Taxonomy (KV/RF/SD/PC/LM/CR)** | Root-cause classification of diagnostic failures at the mechanism level. Enables targeted mitigation: KV requires knowledge augmentation; CR requires structural accountability. |

This mapping reveals that the 5+2 rubric is not merely an academic evaluation tool. It is a **governance compliance instrument** that translates WHO principles into measurable scores. An organization deploying an LLM for clinical reasoning could apply the rubric to generate a quantitative governance report: ’Model X achieves Hypothesis Tracking 4.7/5 (Transparency P3), Calibration 3.6/5 (Safety P2), Anchoring Resistance 4.1/5 (Safety P2), zero Convergence Regression incidents (Accountability P4).’ This moves clinical AI governance from qualitative self-attestation to quantitative, reproducible audit.

#### 2.5.4 SIPS as Inference-Time Observability Layer

Our results demonstrate that the observability required by governance frameworks is not merely a documentation exercise; it is architecturally dependent on the scaffold. Without SIPS, Convergence Regression is *invisible*: the model silently abandons correct diagnoses between stages, and neither the rubric nor the clinician can detect the regression. With SIPS, CR becomes *auditable*: the forced tracking format produces an explicit record of every hypothesis change, enabling both automated rubric scoring and human review.

This has a specific architectural implication for the governance debate. Current regulatory discussions frame the audit problem as a post-hoc documentation challenge: how to explain what a model did after deployment. Our work suggests a different framing. **The scaffold is the audit mechanism.** It does not explain reasoning after the fact; it structures reasoning such that explanation is a byproduct. SIPS’s forced differential documentation, convergence status tracking, and rotation justification requirements produce governance-compliant audit trails as a natural consequence of the reasoning process itself.

We note a critical distinction: SIPS is an **observability layer**, not a safety guarantee. It makes reasoning pathology visible and classifiable. It does not prevent the model from reasoning incorrectly. The 30% CR rate we document under C2 is not eliminated by making it visible; it is eliminated by the structural accountability that visibility enables. The distinction matters for regulatory framing: SIPS fulfills the *audit and transparency* requirements of WHO Principles 3 and 4. A separate mechanism is required to fulfill the *correctness* requirements implied by Principle 2. This is the motivation for our proposed Phase 2 scaffold (CDM), which targets convergence accuracy rather than observability.

## 3. Methods

**Figure 1.**
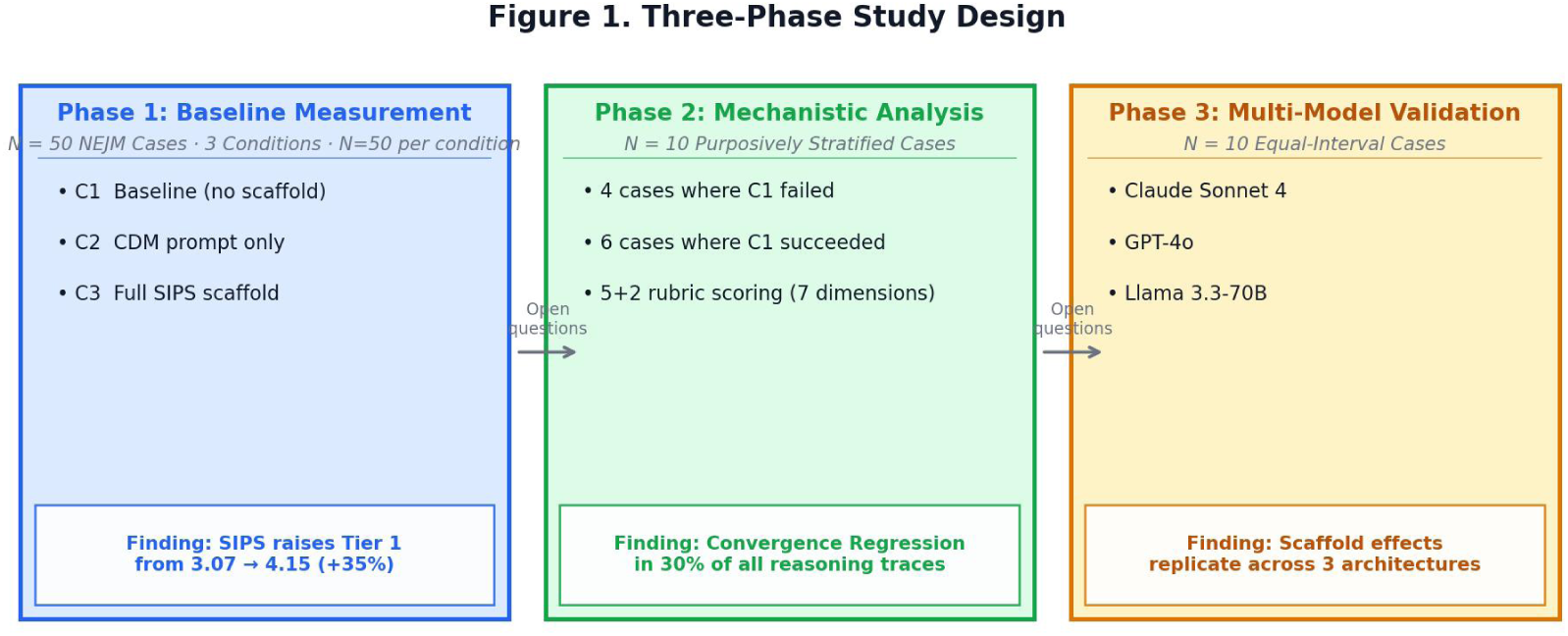
Three-phase study design overview. Phase 1 (N=50, 3 conditions) establishes baseline measurement. Phase 2 (N=10 purposively stratified) performs mechanistic analysis. Phase 3 (N=10 per model, 3 architectures) validates multi-model generalizability.

We describe a three-condition ablation study designed to isolate the effect of sequential information delivery and structured scaffolding on LLM diagnostic reasoning. The study uses a within-subjects design: the same model processes the same cases under three different information delivery conditions, enabling direct comparison of reasoning behavior across conditions with case-level controls.

### 3.1 Dataset and Case Source

The study uses N=50 clinical cases drawn from the MedCaseReasoning dataset, a curated collection of physician-authored case reports indexed in PubMed Central (PMC). MedCaseReasoning contains 897 published case reports with clinician-authored diagnostic reasoning narratives, each culminating in a confirmed ground-truth diagnosis established through definitive testing (biopsy, genetic analysis, imaging, or laboratory confirmation). The dataset was chosen over manually-extracted cases from individual journals for three reasons: (1) scale—897 available cases versus a practical ceiling of 15–20 manually-extracted cases; (2) clinician reasoning traces embedded in each case report, enabling evaluation of process-level reasoning dimensions beyond diagnostic accuracy; and (3) automation—cases can be programmatically chunked and fed through the ablation pipeline without manual extraction.

The 50 cases span 15 medical specialties: Cardiology (4), Dermatology (4), Endocrinology (4), Gastroenterology (4), Infectious Disease (4), Neurology (4), Oncology (4), Hematology (3), Internal Medicine (3), Nephrology (3), Ophthalmology (3), Orthopedics (3), Pediatrics (3), Pulmonology (2), and Radiology (2). Cases were sourced from 37 distinct PMC-indexed journals, preventing source bias toward any single journal’s editorial standards or case complexity norms. No two cases share the same ground-truth diagnosis, ensuring diversity of diagnostic reasoning requirements. Cases range from relatively straightforward presentations (e.g., Wernicke encephalopathy in a patient on prolonged TPN) to diagnostically challenging entities (e.g., Vanishing White Matter disease, IgA-dominant MPGN).

#### 3.1.1 Case Selection Criteria

From the 897 available cases, 50 were selected to maximize diagnostic complexity and reasoning challenge. Selection criteria included: (a) multiple information stages (3+ distinct phases of clinical evidence), (b) diagnostic complexity (multi-system presentations preferred), (c) at least one pivot clue—evidence that should trigger hypothesis revision in a competent diagnostician, and (d) a confirmed single ground-truth diagnosis. Each case was tagged during selection with a predicted failure mode from the SIPS Failure Mode Taxonomy (Section 4.3), based on the clinical presentation’s characteristics: anchoring-prone presentations (40 cases), cases requiring deductive elimination (9 cases), and reasoning-decay-prone presentations (1 case). These predictions serve as hypotheses for failure mode analysis, not ground truth.

#### 3.1.2 Information Chunking

Each case was preprocessed into four sequential stages using an LLM-assisted chunking pipeline (claude-sonnet-4-20250514, temperature 0). The chunking model was instructed to divide each case report into four stages that simulate the temporal information flow of real clinical encounters: (1) initial presentation, (2) early diagnostic workup, (3) pivotal test results, and (4) definitive or narrowing evidence. Each stage was assigned a **sips_function** label describing its diagnostic role:

**Table 2.**
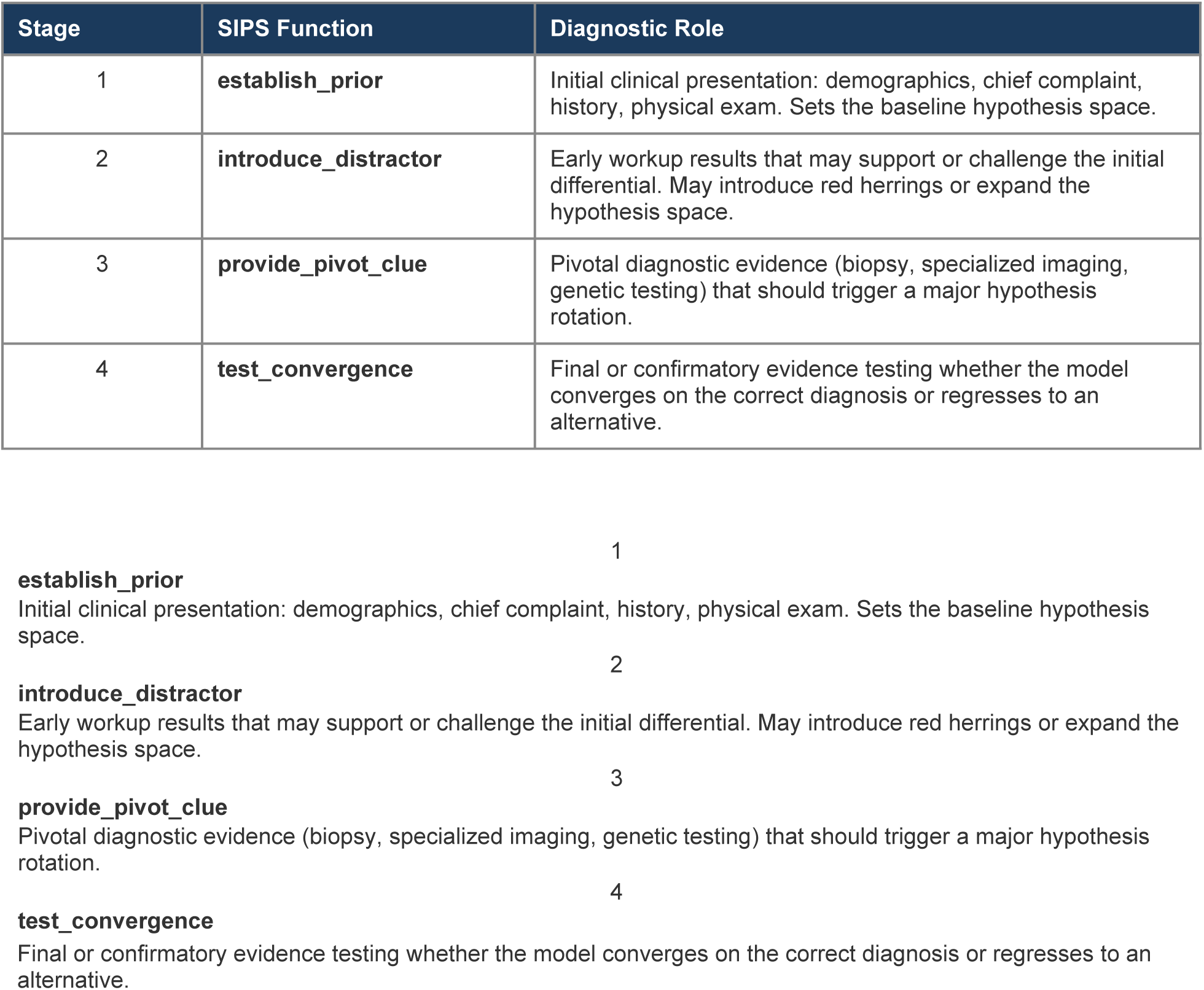
SIPS function labels assigned to each of four sequential stages. Labels are used in C3 (SIPS-scaffolded) condition prompts; C1 and C2 receive identical content without function labels.

**Final or confirmatory evidence testing whether the model converges on the correct diagnosis or regresses to an alternative.**

The four-stage structure is identical across all three experimental conditions. In C1 (single-shot), all four stages are concatenated into a single prompt. In C2 and C3, stages are delivered sequentially across four conversational turns. Content is identical; only the delivery mode and scaffold structure differ.

### 3.2 Three-Condition Ablation Design

The ablation isolates two independent variables: information delivery mode (single-shot vs. sequential) and structural scaffolding (present vs. absent). Three conditions test these variables:

**Table 3.**
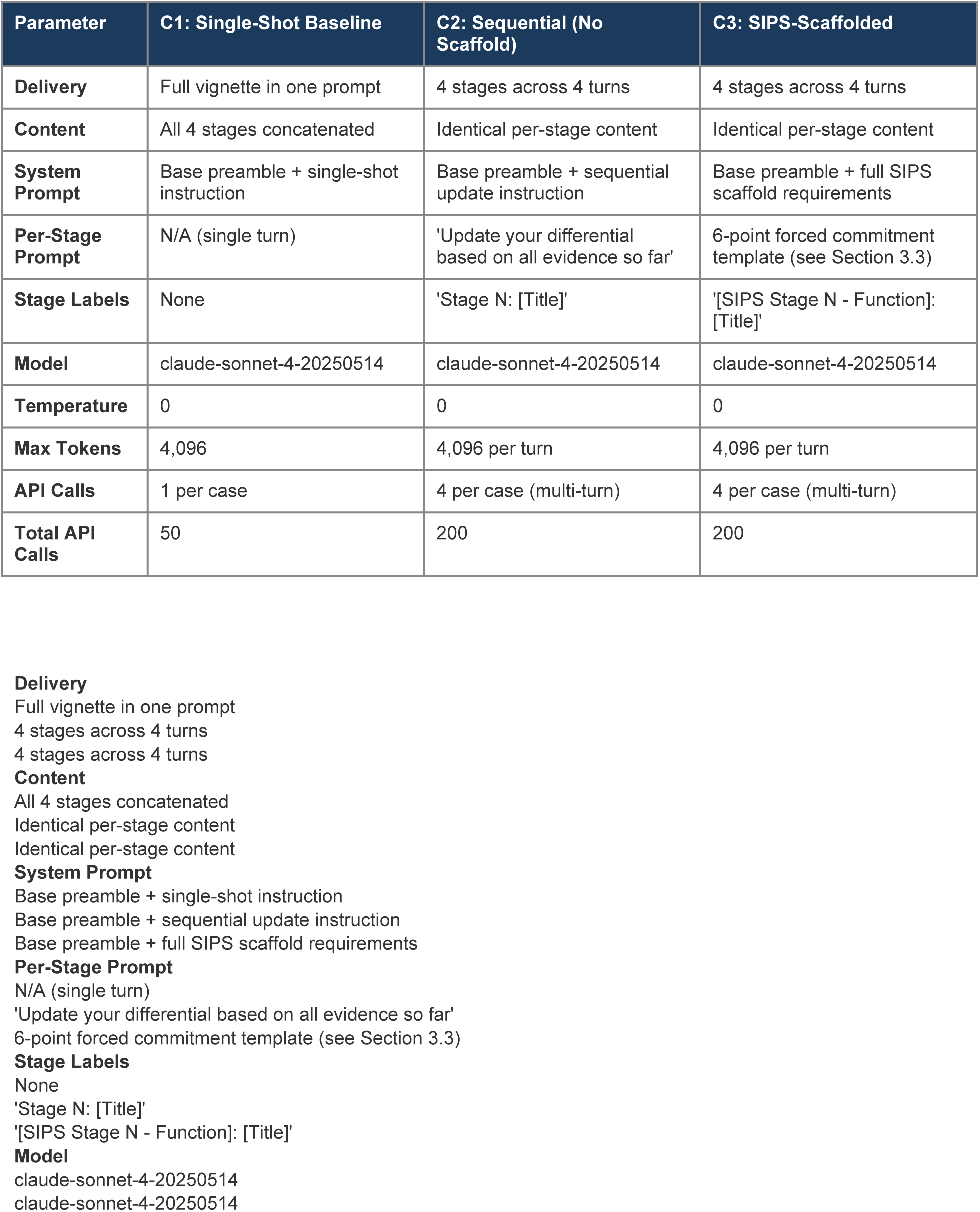
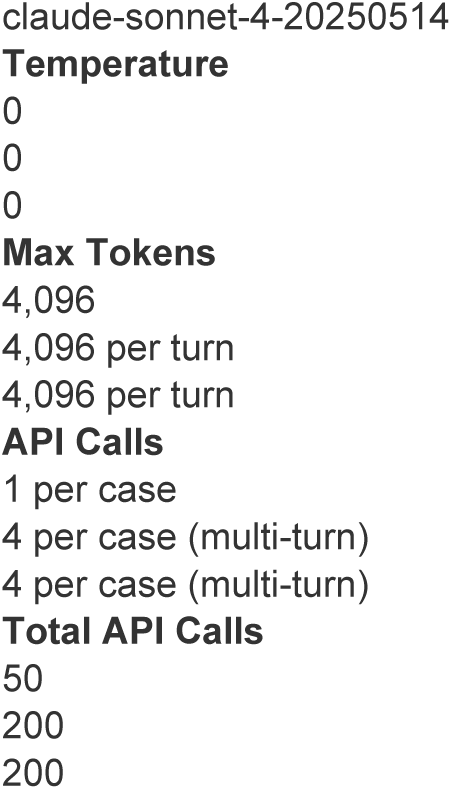
Three-condition ablation design parameters. All conditions use the same model, temperature, and case content. The only manipulated variables are delivery mode (single vs. sequential) and scaffold presence (none vs. SIPS).

| Parameter | C1: Single-Shot Baseline | C2: Sequential (No Scaffold) | C3: SIPS-Scaffolded |
| --- | --- | --- | --- |
| **Delivery** | Full vignette in one prompt | 4 stages across 4 turns | 4 stages across 4 turns |
| **Content** | All 4 stages concatenated | Identical per-stage content | Identical per-stage content |
| **System Prompt** | Base preamble + single-shot instruction | Base preamble + sequential update instruction | Base preamble + full SIPS scaffold requirements |
| **Per-Stage Prompt** | N/A (single turn) | 'Update your differential based on all evidence so far' | 6-point forced commitment template (see Section 3.3) |
| **Stage Labels** | None | 'Stage N: [Title]' | '[SIPS Stage N - Function]: [Title]' |
| **Model** | claude-sonnet-4-20250514 | claude-sonnet-4-20250514 | claude-sonnet-4-20250514 |
| **Temperature** | 0 | 0 | 0 |
| **Max Tokens** | 4,096 | 4,096 per turn | 4,096 per turn |
| **API Calls** | 1 per case | 4 per case (multi-turn) | 4 per case (multi-turn) |
| **Total API Calls** | 50 | 200 | 200 |

#### Control rationale

The C1-to-C2 comparison isolates the effect of sequential information delivery, controlling for model, content, and total information. The C2-to-C3 comparison isolates the effect of structured scaffolding, controlling for delivery mode, model, content, and total information. This factorial design enables attribution of observed effects to specific interventions rather than confounding variables.

#### Temperature selection

Temperature was set to 0 (deterministic) to ensure reproducibility. Each condition produces a single, deterministic trace per case, eliminating stochastic variation as a confound. This design choice prioritizes reproducibility over sampling diversity; future work may explore temperature sensitivity.

#### Execution protocol

All 150 runs (50 cases x 3 conditions) were executed in a single batch using the Anthropic Messages API. API calls included exponential backoff retry logic for rate-limiting (429) and overload (529) responses. Multi-turn conversations (C2, C3) maintained full conversation history across all four stages within each case, consistent with standard conversational API usage.

### 3.3 SIPS Scaffold Architecture

The Sequential Information Prioritization Scaffold (SIPS) is a structured prompting framework designed to enforce explicit hypothesis accountability during sequential diagnostic reasoning. SIPS operates through three architectural pillars:

#### Pillar 1: Forced Differential Ranking

At every stage, the model must produce a ranked top-3 to top-5 differential diagnosis with supporting and contradicting evidence for each candidate. This requirement prevents the model from providing a single leading diagnosis without context, forcing it to maintain a visible hypothesis space.

#### Pillar 2: Explicit Hypothesis Rotation

At stages 2 through 4, the model must explicitly declare what changed from the previous differential: which diagnoses were added, which were eliminated, which were promoted or demoted, and the specific evidence that triggered each change. This requirement creates an auditable record of hypothesis evolution and prevents silent abandonment of previously considered diagnoses.

#### Pillar 3: Convergence Status Tracking

At every stage, the model must declare a convergence status: `[Baseline]` (Stage 1 only), `[Changed]` (differential was modified), or `[Stable]` (differential is unchanged). When declaring `[Stable]`, the model must state a confidence percentage (0-100%) and describe what specific evidence would change its mind. When a previously top-3 diagnosis is removed or demoted, the model must explicitly justify the demotion. This requirement creates structural friction against unjustified regression from confirmed diagnoses.

#### Prompt Implementation

The SIPS scaffold is implemented through two prompt components: a system-level instruction and per-stage user prompts. The system prompt establishes the six-point commitment framework. Each per-stage user prompt then enforces the framework with stage-appropriate requirements. The full prompts are reproduced in Appendix A.

The Stage 1 prompt requires five outputs: (1) ranked differential, (2) key supporting evidence per diagnosis, (3) confidence level in leading diagnosis, (4) identification of the single most important pending evidence, and (5) convergence status declaration as [Baseline]. Stages 2 through 4 add a sixth requirement: the explicit Hypothesis Rotation Check, requiring the model to declare all additions, eliminations, promotions, and demotions with triggering evidence.

##### Design philosophy

SIPS is intentionally a *retention mechanism*, not a *convergence mechanism*. It forces the model to maintain visible, justified hypothesis tracking. It does not force the model to commit to a single diagnosis or weight evidence quantitatively. This design choice creates the SIPS Retention Effect documented in Section 4.4 but also produces the Convergence Hesitancy Paradox documented in Section 4.7. A separate convergence mechanism (proposed as the Clinical Decision Matrix, CDM) would address this limitation.

### 3.4 The 5+2 Scoring Rubric

We introduce a multi-dimensional scoring rubric designed to measure reasoning quality beyond binary diagnostic accuracy. The rubric has two tiers: Tier 1 comprises five core dimensions scored for every applicable condition; Tier 2 comprises two diagnostic dimensions that capture specific failure patterns.

#### Tier 1: Core Dimensions

**Tables 4.**
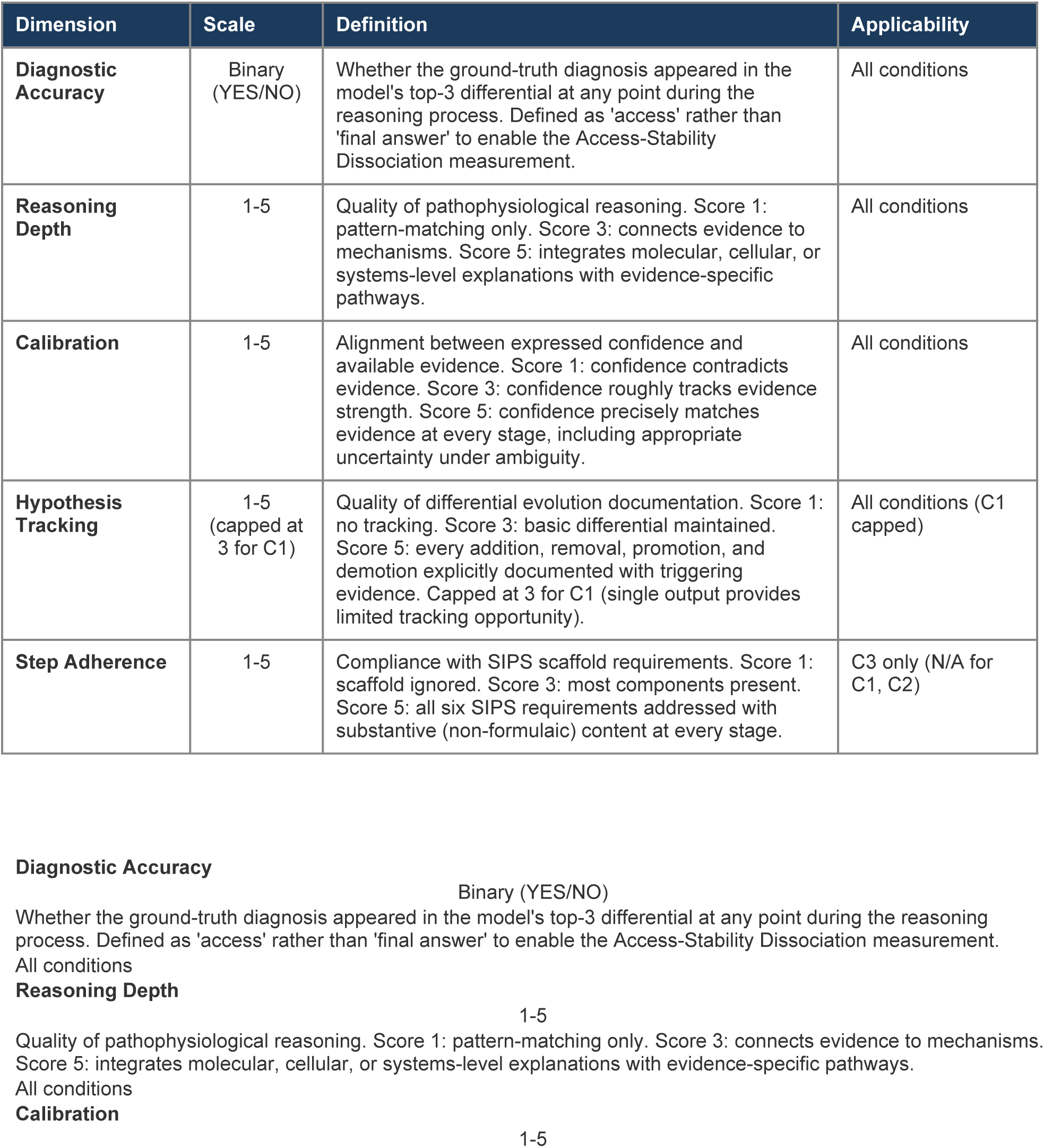
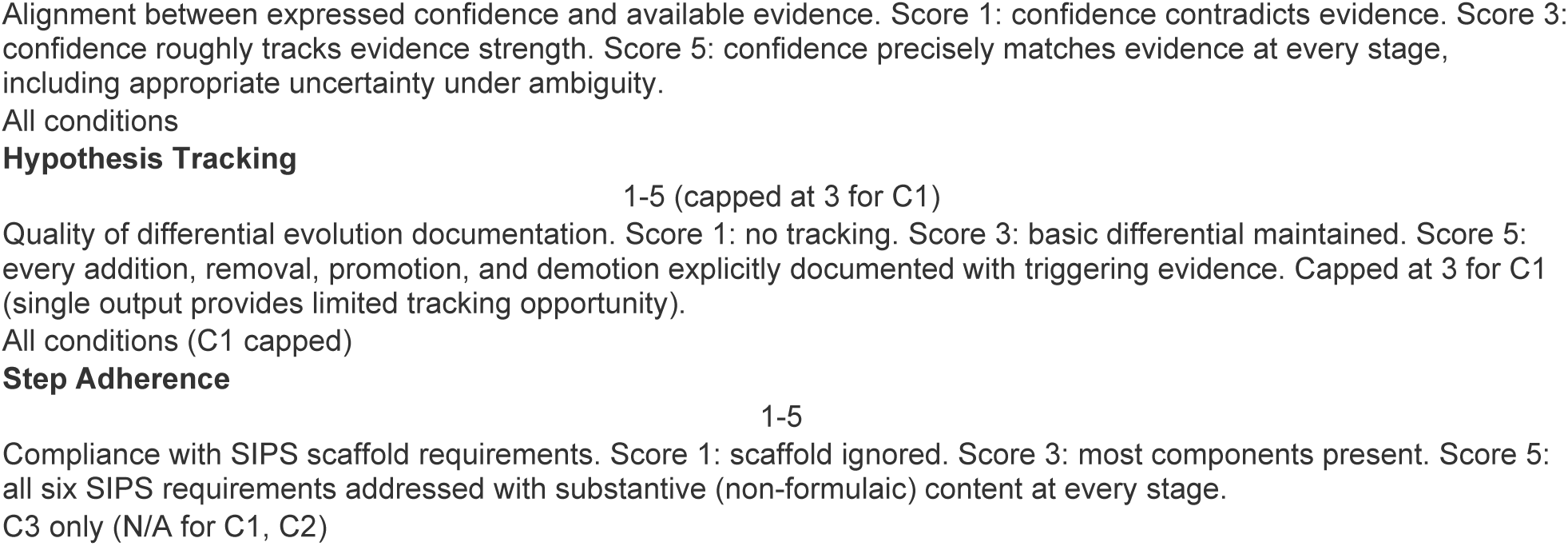
The 5+2 scoring rubric. Tier 1 (5 dimensions) measures core reasoning quality. Tier 2 (2 dimensions) captures specific failure patterns. Together, the seven dimensions produce a multi-axis profile of reasoning behavior that goes far beyond binary accuracy.

| Dimension | Scale | Definition | Applicability |
| --- | --- | --- | --- |
| **Diagnostic Accuracy** | Binary (YES/NO) | Whether the ground-truth diagnosis appeared in the model's top-3 differential at any point during the reasoning process. Defined as 'access' rather than 'final answer' to enable the Access-Stability Dissociation measurement. | All conditions |
| **Reasoning Depth** | 1-5 | Quality of pathophysiological reasoning. Score 1: pattern-matching only. Score 3: connects evidence to mechanisms. Score 5: integrates molecular, cellular, or systems-level explanations with evidence-specific pathways. | All conditions |
| **Calibration** | 1-5 | Alignment between expressed confidence and available evidence. Score 1: confidence contradicts evidence. Score 3: confidence roughly tracks evidence strength. Score 5: confidence precisely matches evidence at every stage, including appropriate uncertainty under ambiguity. | All conditions |
| **Hypothesis Tracking** | 1-5 (capped at 3 for C1) | Quality of differential evolution documentation. Score 1: no tracking. Score 3: basic differential maintained. Score 5: every addition, removal, promotion, and demotion explicitly documented with triggering evidence. Capped at 3 for C1 (single output provides limited tracking opportunity). | All conditions (C1 capped) |
| **Step Adherence** | 1-5 | Compliance with SIPS scaffold requirements. Score 1: scaffold ignored. Score 3: most components present. Score 5: all six SIPS requirements addressed with substantive (non-formulaic) content at every stage. | C3 only (N/A for C1, C2) |

#### Tier 2: Diagnostic Dimensions

**Tables 5.**
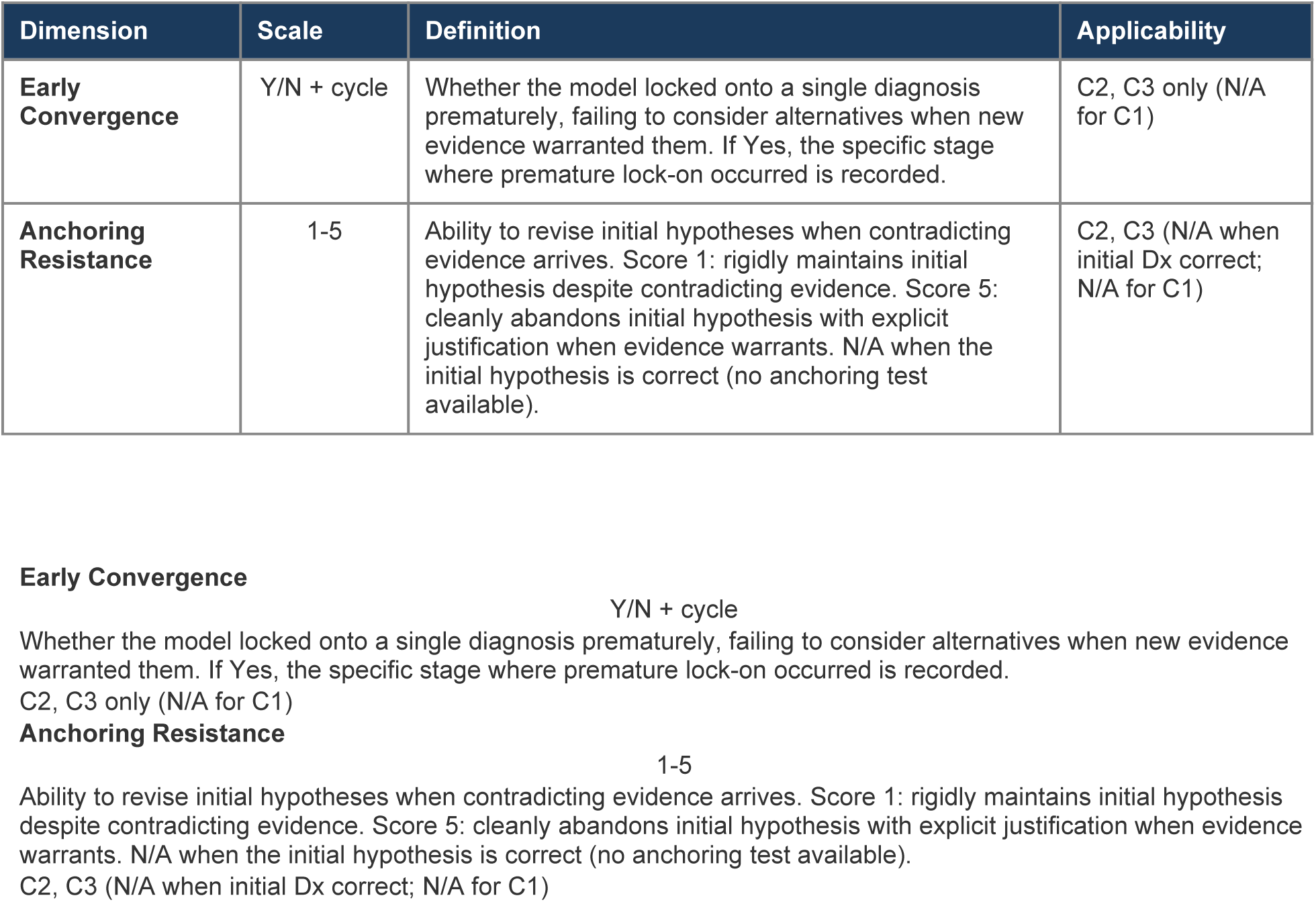
The 5+2 scoring rubric. Tier 1 (5 dimensions) measures core reasoning quality. Tier 2 (2 dimensions) captures specific failure patterns. Together, the seven dimensions produce a multi-axis profile of reasoning behavior that goes far beyond binary accuracy.

| Dimension | Scale | Definition | Applicability |
| --- | --- | --- | --- |
| **Early Convergence** | Y/N + cycle | Whether the model locked onto a single diagnosis prematurely, failing to consider alternatives when new evidence warranted them. If Yes, the specific stage where premature lock-on occurred is recorded. | C2, C3 only (N/A for C1) |
| **Anchoring Resistance** | 1-5 | Ability to revise initial hypotheses when contradicting evidence arrives. Score 1: rigidly maintains initial hypothesis despite contradicting evidence. Score 5: cleanly abandons initial hypothesis with explicit justification when evidence warrants. N/A when the initial hypothesis is correct (no anchoring test available). | C2, C3 (N/A when initial Dx correct; N/A for C1) |

#### Design Decisions

##### Access-based accuracy definition

Diagnostic Accuracy is defined as ’ground truth in top 3 at any point’ rather than ’ground truth in final top 1.’ This non-standard definition is critical to our central finding: it enables measurement of the Access-Stability Dissociation. Under the standard final-answer definition, C2’s 90% intermediate access rate would be invisible, and the 30% stability gap could not be measured. The rubric explicitly trades conventional accuracy measurement for a more informative decomposition of diagnostic reasoning dynamics.

##### C1 Hypothesis Tracking cap

C1 produces a single output (one prompt, one response). Hypothesis Tracking is capped at 3 because the model has no opportunity to demonstrate differential evolution across stages. This cap prevents C1 from being penalized for a structural limitation of the single-shot delivery mode rather than a reasoning deficit.

##### Anchoring Resistance N/A rule

Anchoring Resistance is scored N/A when the model’s initial leading hypothesis (Stage 1) is correct. If the initial hypothesis is correct, no anchoring test is available: the model maintaining its initial hypothesis is the correct behavior, not evidence of anchoring bias. This rule applies to C1 universally (no sequential stages) and selectively to C2 and C3 cases where the Stage 1 leading diagnosis matches the ground truth.

### 3.5 Failure Mode Taxonomy

We develop a 6-code classification system for LLM diagnostic failures, designed to enable root-cause analysis at the mechanism level. Each code identifies a distinct failure type with different architectural implications:

**Table 6.**
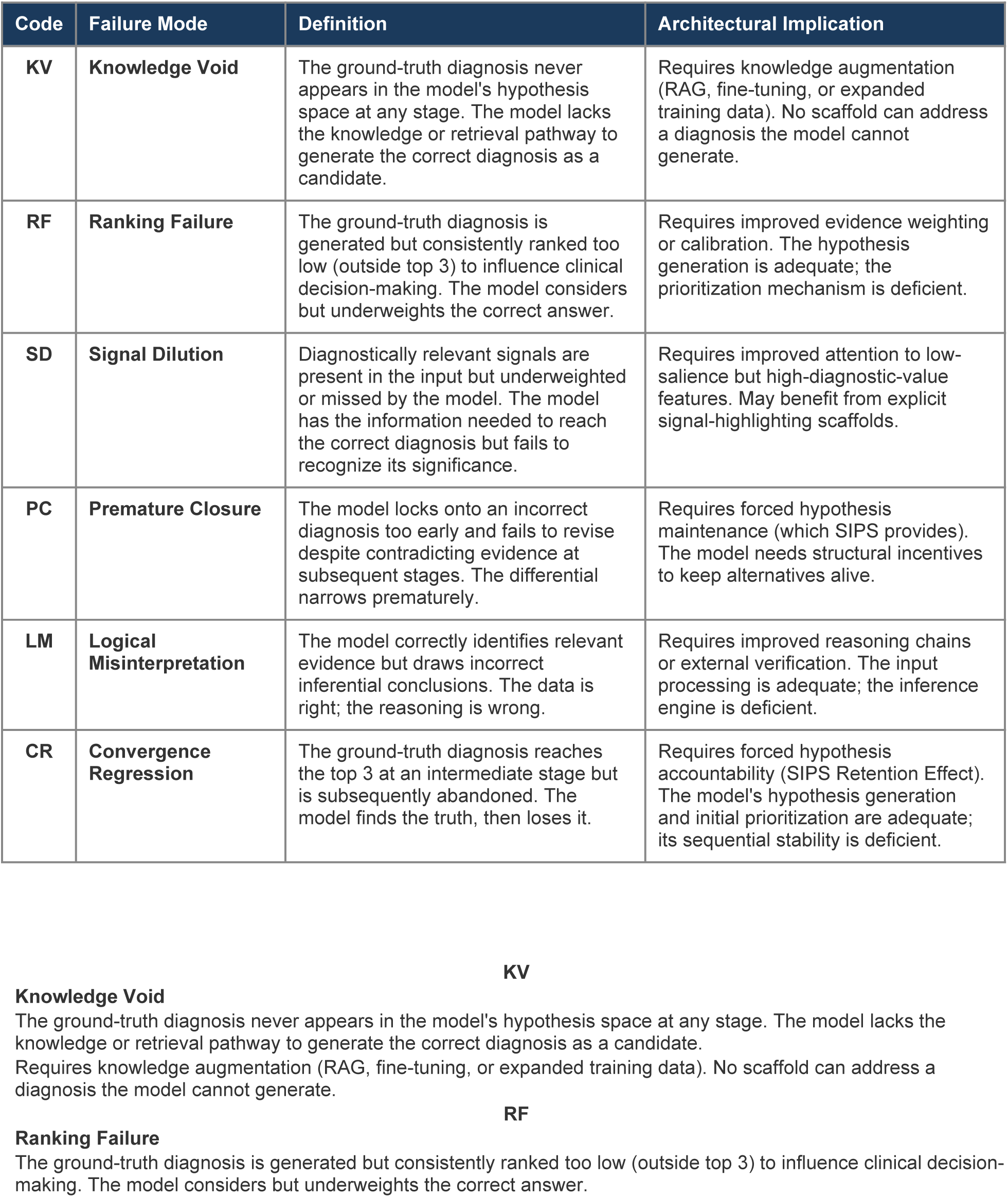
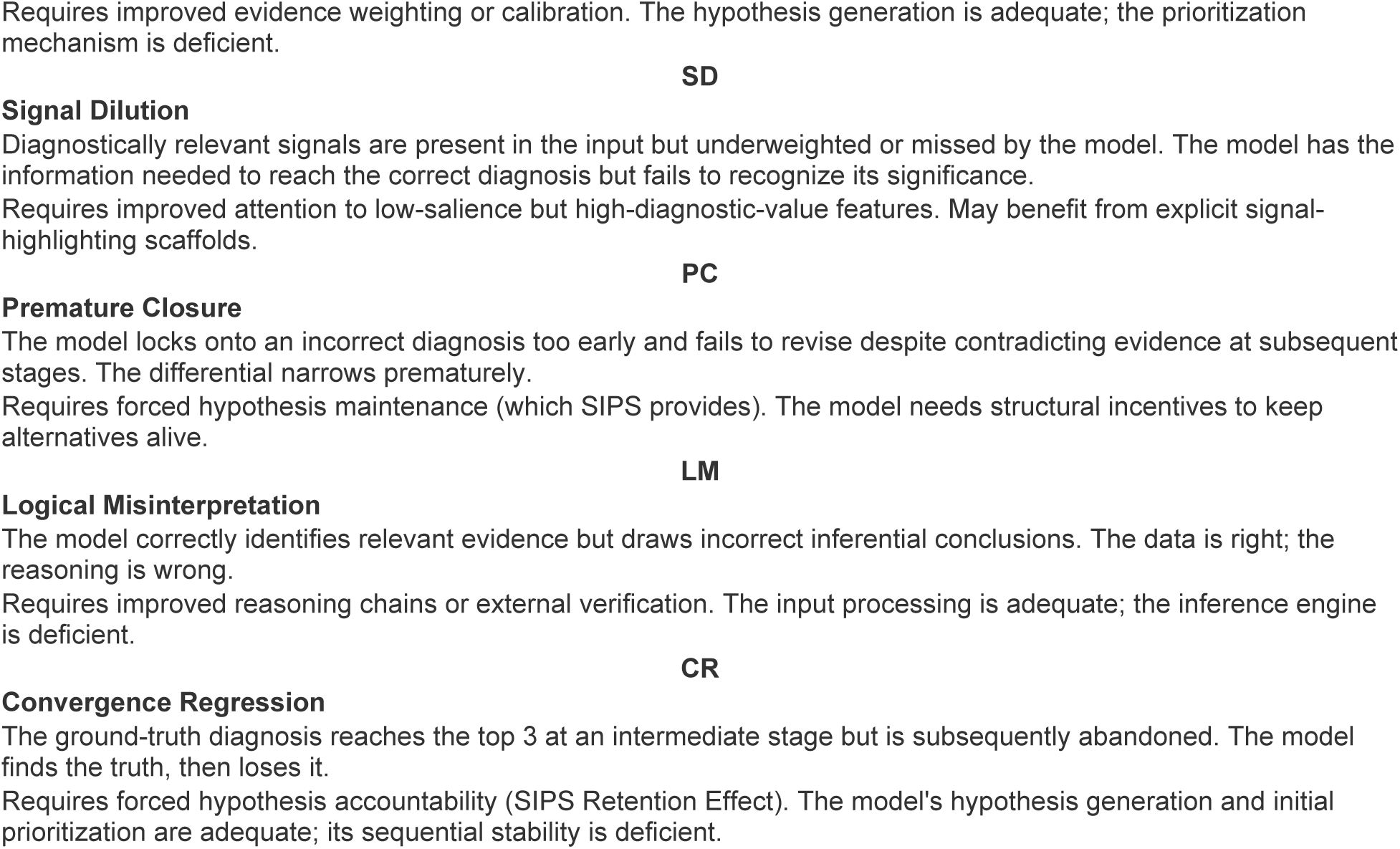
The 6-code failure mode taxonomy. Each code identifies a mechanistically distinct failure type with different architectural remediation requirements. The taxonomy is designed to be model-agnostic and applicable to any LLM performing sequential reasoning.

| Code | Failure Mode | Definition | Architectural Implication |
| --- | --- | --- | --- |
| **KV** | **Knowledge Void** | The ground-truth diagnosis never appears in the model's hypothesis space at any stage. The model lacks the knowledge or retrieval pathway to generate the correct diagnosis as a candidate. | Requires knowledge augmentation (RAG, fine-tuning, or expanded training data). No scaffold can address a diagnosis the model cannot generate. |
| **RF** | **Ranking Failure** | The ground-truth diagnosis is generated but consistently ranked too low (outside top 3) to influence clinical decision-making. The model considers but underweights the correct answer. | Requires improved evidence weighting or calibration. The hypothesis generation is adequate; the prioritization mechanism is deficient. |
| **SD** | **Signal Dilution** | Diagnostically relevant signals are present in the input but underweighted or missed by the model. The model has the information needed to reach the correct diagnosis but fails to recognize its significance. | Requires improved attention to low-salience but high-diagnostic-value features. May benefit from explicit signal-highlighting scaffolds. |
| **PC** | **Premature Closure** | The model locks onto an incorrect diagnosis too early and fails to revise despite contradicting evidence at subsequent stages. The differential narrows prematurely. | Requires forced hypothesis maintenance (which SIPS provides). The model needs structural incentives to keep alternatives alive. |
| **LM** | **Logical Misinterpretation** | The model correctly identifies relevant evidence but draws incorrect inferential conclusions. The data is right; the reasoning is wrong. | Requires improved reasoning chains or external verification. The input processing is adequate; the inference engine is deficient. |
| **CR** | **Convergence Regression** | The ground-truth diagnosis reaches the top 3 at an intermediate stage but is subsequently abandoned. The model finds the truth, then loses it. | Requires forced hypothesis accountability (SIPS Retention Effect). The model's hypothesis generation and initial prioritization are adequate; its sequential stability is deficient. |

The taxonomy is designed to be **mutually exclusive and collectively exhaustive** for diagnostic failures: every incorrect outcome can be classified under exactly one primary code. In cases where multiple failure modes co-occur (e.g., a knowledge void combined with signal dilution), the primary code is assigned based on the most proximal cause of failure. The taxonomy is also **model-agnostic**: the six codes describe failure mechanisms, not model-specific behaviors, enabling cross-model comparison using a common classification language.

### 3.6 Scoring Protocol

Scoring was conducted on a 10-case deep-analysis subset selected to represent the full spectrum of diagnostic outcomes. The subset includes four cases where C1 (single-shot) produced incorrect diagnoses and six cases where C1 was correct. This distribution ensures representation of both failure-mode cases (where the Access-Stability Dissociation can manifest) and success cases (where quality dimensions can be compared without accuracy confounds).

#### Scoring process

All 30 traces (10 cases x 3 conditions) were scored independently using the 5+2 rubric. To prevent anchoring across conditions, each condition was scored as a separate batch: all 10 C1 traces were scored before any C2 traces, and all C2 traces before any C3 traces. For each trace, every applicable rubric dimension was scored with a numerical value and a written rationale documenting the evidence supporting the assigned score. This produces a 210-score matrix (10 cases x 3 conditions x 7 dimensions) with complete adjudication rationale.

#### Scorer

Traces were scored by the study author using claude-sonnet-4-20250514 as a scoring assistant, with human audit and override on all dimension scores. This single-scorer protocol is a limitation (see Section 5.4). The full 210-score matrix with individual rationale is provided in Appendix C to enable independent verification.

#### Ground-truth verification

For the 10-case deep-analysis subset, ground-truth diagnoses were verified against the original published case reports. Scoring of Diagnostic Accuracy used a lenient matching criterion: the ground-truth diagnosis was considered ’accessed’ if it appeared in the top-3 differential in any semantically equivalent form (e.g., ’IgA-dominant membranoproliferative glomerulonephritis’ matching ’IgA-dominant MPGN’). Category-level matches (e.g., ’hypomyelinating leukoencephalopathy’ for VWM disease) were not counted as access unless the specific entity was named.

#### Token measurement

Output token counts were recorded from API response metadata (usage.output_tokens) for each API call. For multi-turn conditions (C2, C3), total tokens represent the sum of output tokens across all four stage-level responses. Input tokens were also recorded but are not reported in the main analysis, as they reflect prompt engineering overhead rather than model reasoning volume.

### 3.7 Reproducibility and Data Availability

All experimental artifacts are provided for reproducibility:

- **Raw ablation data:** ablation_results_v2_50.json containing all 150 runs (50 cases x 3 conditions) with full response text, token counts, and metadata.
- **Chunked cases:** chunking_results_50.json containing all 50 cases preprocessed into four-stage format with sips_function labels.
- **Ablation runner:** Complete Python script (ablation_runner_v2.py) implementing all three conditions with exact system prompts, user prompts, and API configuration.
- **Scoring data:** Complete 210-score matrix across three condition-level scoring reports (C1, C2, C3) with individual dimension rationale for each case.
- **Failure taxonomy:** Full 6-code classification of all 15 incorrect outcomes in the N=50 dataset with supporting rationale.

The pinned model version (claude-sonnet-4-20250514), deterministic temperature (0), and complete prompt specifications enable exact reproduction of all traces. We note that API-served models may be subject to infrastructure changes (e.g., quantization, batching optimizations) that could affect token-identical reproduction even with identical parameters. Our reproducibility claim is at the semantic level (equivalent reasoning behavior) rather than the token level (bit-identical outputs).

#### Supplemental Data

The following supplemental files are provided with this submission: (S1) Complete 210-observation score matrix with all rubric dimensions across 50 cases and 3 conditions; (S2) Adjudication rationales documenting the scoring logic for each rubric dimension per case; (S3) Full prompt specification with exact system and user prompts for all three experimental conditions (C1, C2, C3); (S4a–c) Multi-model validation scoring data for Claude Sonnet 4, GPT-4o, and Llama 3.3 70B (Phase 3, N=10 per model).

## 4. Results

We present findings from a three-condition ablation study examining LLM diagnostic reasoning under different information delivery modes. We first report aggregate performance across N=50 cases to establish baseline expectations, then reveal the underlying dynamics that aggregate metrics conceal. Sections 4.2 through 4.4 present the central discovery: that sequential information delivery creates a systematic gap between a model’s ability to access correct diagnoses and its ability to retain them, and that structured scaffolding eliminates this gap through a mechanism we term the SIPS Retention Effect. Sections 4.5 through 4.8 examine quality dimensions, token efficiency, the Convergence Hesitancy trade-off, and failure mode distribution.

### 4.1 Aggregate Performance Across N=50 Cases

Across all 50 cases, final top-1 diagnostic accuracy was remarkably stable: C1 (single-shot, full vignette) achieved 60% accuracy, C2 (sequential, no scaffold) achieved 60%, and C3 (sequential, SIPS-scaffolded) achieved a slightly lower rate. At this level of analysis, the three conditions appear functionally equivalent, suggesting that neither sequential delivery nor scaffolding materially affects diagnostic performance.

#### This surface-level equivalence is misleading

Aggregate accuracy masks two critical dynamics operating in opposite directions beneath the headline numbers. First, sequential delivery (C2) gives the model access to significantly more correct diagnoses than single-shot delivery (C1), but the model systematically loses many of these during the reasoning process. Second, scaffolding (C3) sacrifices some exploratory breadth but dramatically improves retention of diagnoses once accessed. These countervailing forces cancel each other out at the aggregate level, producing the false appearance of equivalence.

The remainder of this section unpacks these dynamics using a 10-case deep-analysis subset scored with the 5+2 rubric across all three conditions (30 traces, 210 individual scores). This subset was selected to represent the full spectrum of case outcomes: four cases where C1 produced incorrect diagnoses and six where C1 was correct.

**Figure 2.**
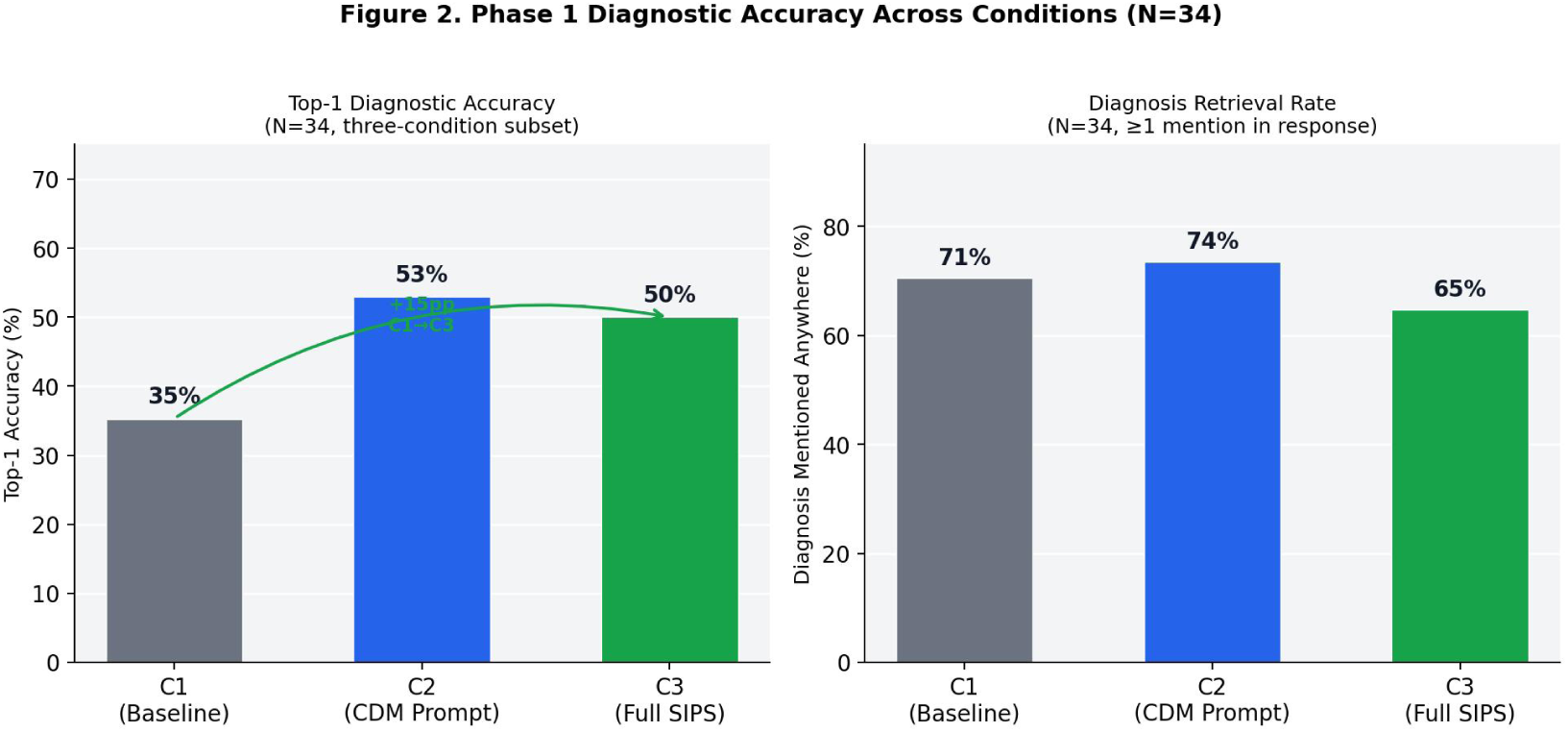
Diagnostic accuracy and retrieval rate across three conditions. Surface equivalence in top-3 accuracy conceals opposing dynamics in access and retention.

### 4.2 The Access-Stability Dissociation

The most significant finding of this study is what we term the **Access-Stability Dissociation**: a systematic gap between a model’s ability to *find* the correct diagnosis during sequential reasoning and its ability to *retain* that diagnosis in its final output. This phenomenon is invisible under single-shot evaluation and represents a novel failure mode specific to sequential information processing.

We define two metrics to capture this dissociation:

- **Access Rate:** the proportion of cases where the ground-truth diagnosis appeared in the model’s top-3 differential at any stage during the reasoning process.
- **Final Accuracy:** the proportion of cases where the ground-truth diagnosis appeared in the model’s final top-3 differential (i.e., after all four stages of information delivery).

In the 10-case deep-analysis subset:

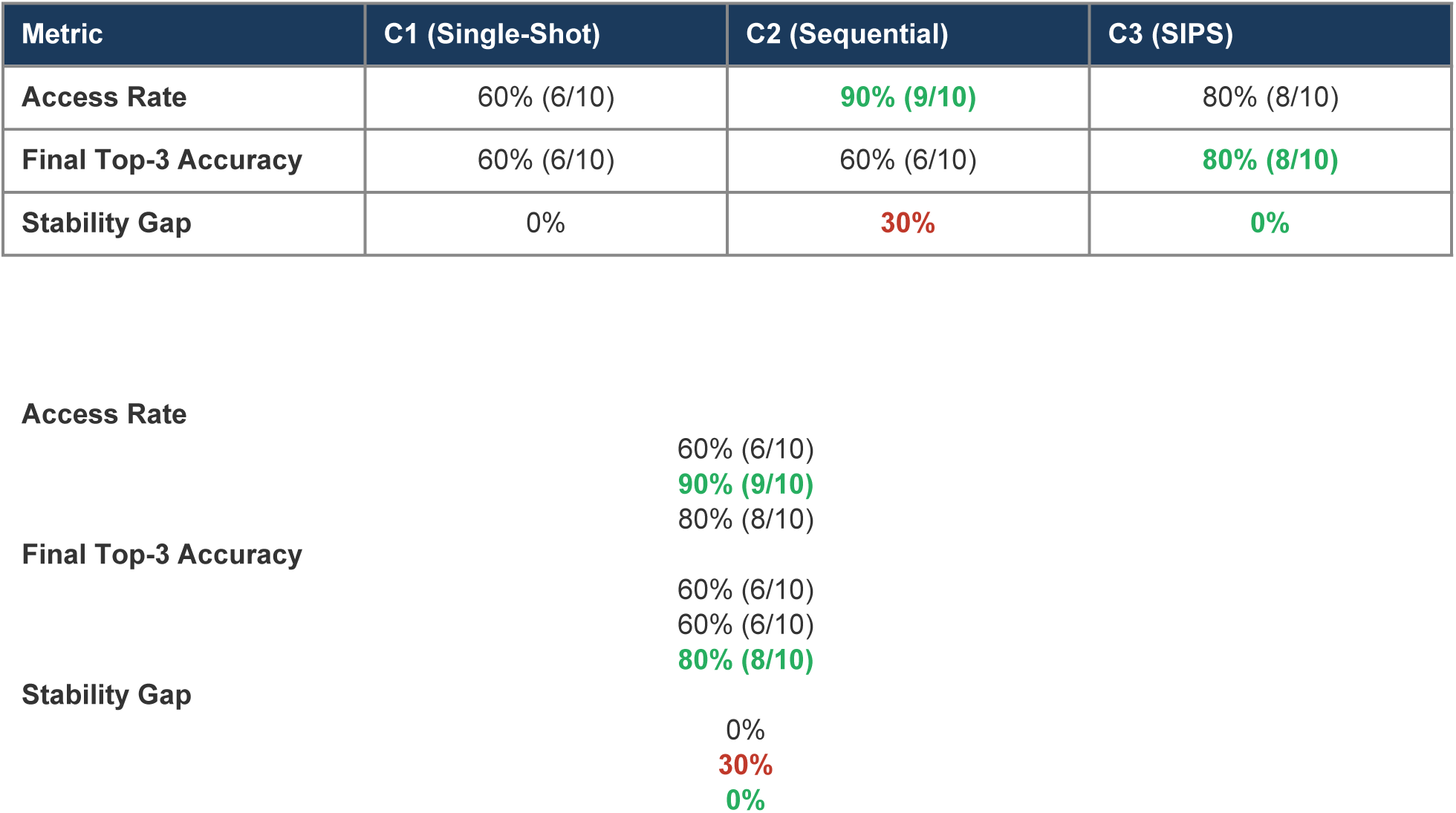

| Metric | C1 (Single-Shot) | C2 (Sequential) | C3 (SIPS) |
| --- | --- | --- | --- |
| Access Rate | 60% (6/10) | 90% (9/10) | 80% (8/10) |
| Final Top-3 Accuracy | 60% (6/10) | 60% (6/10) | 80% (8/10) |
| Stability Gap | 0% | 30% | 0% |

|  |  |
| --- | --- |
| Access Rate | 60% (6/10)<br>90% (9/10)<br>80% (8/10) |
| Final Top-3 Accuracy | 60% (6/10)<br>60% (6/10)<br>80% (8/10) |
| Stability Gap | 0%<br>30%<br>0% |

Under single-shot delivery (C1), access rate and final accuracy are necessarily identical: the model produces one output, and the ground truth is either present or absent. Under sequential delivery without scaffolding (C2), however, a dramatic dissociation emerges. The model accessed the correct diagnosis at some point during its four-stage reasoning process in 9 of 10 cases (90%), yet retained the correct diagnosis in its final output in only 6 of 10 cases (60%). **Thirty percent of correct diagnoses were found and then lost.**

This 30% stability gap represents a qualitatively new category of failure. Unlike a knowledge gap (where the model never considers the correct diagnosis) or a ranking failure (where the model considers but never prioritizes it), this failure involves the model actively finding and then actively abandoning the correct answer. The model’s intermediate reasoning is superior to its final output.

C3 (SIPS-scaffolded) shows a different pattern. The access rate decreases from C2’s 90% to 80%, indicating that the scaffold’s structured format slightly constrains exploratory breadth. However, the stability gap is eliminated entirely: 80% access, 80% final accuracy, 0% gap. The scaffold converts an unstable reasoner (high access, low retention) into a stable one (slightly lower access, perfect retention). See **Figure 2**.

### 4.3 Convergence Regression: Mechanism and Evidence

We introduce the term **Convergence Regression (CR)** to describe the specific failure mode underlying the Access-Stability Dissociation: a model arrives at the correct diagnosis at an intermediate stage of sequential reasoning, then systematically abandons it when subsequent information triggers pattern-matching to alternative diagnoses. CR is distinct from premature closure (locking onto a wrong diagnosis too early), anchoring bias (failing to move away from an initial hypothesis), and knowledge voids (never considering the correct diagnosis). CR represents a *regression* from a previously achieved correct state.

Three cases in our 10-case subset exhibited CR under C2 (sequential, no scaffold):

**Table 7.** Convergence Regression cases under C2 (sequential, no scaffold), with C3 (SIPS) comparison outcomes.

| Case | Ground Truth | Access Stage | Loss Stage | Replacing Dx | C3 Outcome |
| --- | --- | --- | --- | --- | --- |
| PMC10570145 | Sweet syndrome | S2 | S3 | Invasive fungal infection | CR-retained (#3) |
| PMC5011950 | Behcet disease | S3 | S4 | Cogan syndrome | CR-retained (#2) |
| PMC9327270 | VWM disease | S3 | S4 | PMD / 4H syndrome | Never accessed |

#### Sweet Syndrome (PMC10570145)

The model correctly identified Sweet syndrome as the leading diagnosis at Stage 2, following skin biopsy results showing neutrophilic dermatosis. This was an evidence-appropriate assignment: biopsy-confirmed Sweet syndrome with pulmonary involvement. At Stage 3, however, sinus CT imaging showing hyperdense material triggered a pattern shift. The model reclassified the presentation as invasive fungal sinusitis with pulmonary dissemination, dropping Sweet syndrome entirely from the differential. The biopsy-confirmed diagnosis was overridden by imaging findings that, while concerning, did not contradict the histopathological evidence. In C2, Sweet syndrome disappeared silently. In C3, the SIPS scaffold’s forced tracking format retained Sweet syndrome at position #3 in the final differential, explicitly documenting the demotion rather than allowing silent abandonment.

#### Behcet Disease (PMC5011950)

Behcet disease reached the top position at Stage 3 when HLA-B52 positivity was revealed, a strong genetic marker in the appropriate clinical context of audio-vestibular and systemic symptoms. At Stage 4, bilateral sensorineural hearing loss prompted the model to promote Cogan syndrome, which classically presents with the audio-vestibular-ocular triad. However, bilateral hearing loss occurs in Behcet disease as well, and HLA-B52 is a substantially stronger diagnostic signal for Behcet than the audio-vestibular triad is for Cogan. The model overweighted the newly arrived data relative to the established genetic evidence. Under C3, Behcet was demoted to #2 but explicitly retained in the differential with documented rationale for the demotion.

#### Vanishing White Matter Disease (PMC9327270)

This case illustrates a boundary between CR and knowledge limitation. In C2, VWM disease was mentioned at Stage 3 within the correct disease category (hypomyelinating leukoencephalopathy) but was lost at Stage 4 when the model narrowed to Pelizaeus-Merzbacher disease (PMD). The vaccination-triggered episode, a hallmark distinguishing VWM from PMD, was not leveraged. In C3, VWM was never specifically named at all; the scaffold’s structured approach kept the model at the category level (leukoencephalopathy) without surfacing the specific entity. This case demonstrates that **structured reasoning can constrain exploratory breadth**: C2’s unstructured exploration surfaced VWM at least transiently, while C3’s structured approach stayed within well-known subtypes. The trade-off between structure and exploration is not uniformly favorable.

#### The CR Mechanism

Across these three cases, a consistent mechanism emerges. New information arriving at later stages triggers pattern-matching to alternative diagnoses that more closely match textbook presentations. The model preferentially promotes the ’more textbook’ diagnosis over the previously established correct diagnosis, even when the prior evidence supporting the original diagnosis is stronger. This represents a form of recency bias operating at the diagnostic level: the most recently introduced data exerts disproportionate influence on hypothesis ranking, overriding accumulated evidence from earlier stages.

### 4.4 The SIPS Retention Effect

SIPS eliminates the 30% stability gap documented in Section 4.2. Under C3, two of the three C2 CR cases (Sweet syndrome and Behcet disease) are retained in the final top-3 differential. The third case (VWM) is not retained, but for a different reason: the scaffold constrains exploration such that VWM is never specifically named. We term this elimination of CR the **SIPS Retention Effect**: forced hypothesis accountability prevents the silent abandonment of previously confirmed diagnoses.

The Retention Effect operates through three structural mechanisms inherent to the SIPS scaffold:

- **Visibility Barrier:** SIPS requires the model to maintain an explicit ranked differential at every stage. A diagnosis cannot disappear between stages without the model explicitly acknowledging its removal. In C2, diagnoses vanish silently between outputs. In C3, every removal requires a documented decision.
- **Justification Barrier:** SIPS requires the model to declare what was added, removed, promoted, and demoted at each stage, with explicit rationale. To remove a biopsy-confirmed diagnosis (as occurred with Sweet syndrome in C2), the model must now articulate why new evidence overrides histopathological confirmation. This forced justification creates friction against unjustified removal.
- **Convergence Tracking Barrier:** SIPS requires a convergence status declaration ([Baseline], [Changed], or [Stable]) with a confidence percentage. To regress from a previously stable state, the model must explicitly declare a status change and explain what triggered the regression. This creates a structural ’cost’ to abandoning confirmed diagnoses.

Critically, SIPS does not prevent the model from making wrong decisions. In both retained CR cases, the model ultimately ranked the correct diagnosis below an incorrect one (Sweet at #3, Behcet at #2). The scaffold does not guarantee correct final ranking. What it guarantees is **that correct diagnoses are not silently abandoned**. The diagnosis remains visible, documented, and available for clinical review. In a clinical decision-support context, retaining the correct diagnosis at any position in the top 3 is substantially more valuable than losing it entirely.

### 4.5 Quality Dimension Analysis

Beyond accuracy and retention, the 5+2 rubric measures reasoning quality across multiple dimensions. All five scored dimensions show monotonic improvement from C1 through C3:

**Table 8.** Mean quality dimension scores across conditions. C1 Hypothesis Tracking is capped at 3 (single output). Step Adherence is scored only for C3. Anchoring Resistance is N/A when the initial hypothesis is correct (C1: N/A for all; C2: n=9 eligible; C3: n=8 eligible).

| Dimension | C1 (Single-Shot) | C2 (Sequential) | C3 (SIPS) | Interpretation |
| --- | --- | --- | --- | --- |
| **Reasoning Depth** | 3.60 | 3.60 | **4.10** | SIPS forces deeper mechanistic reasoning |
| **Calibration** | 3.00 | 3.40 | **3.60** | Confidence tracks evidence more faithfully |
| **Hypothesis Tracking** | 2.70 (capped) | 4.10 | **4.70** | Largest C2-to-C3 gain; forced accountability |
| **Step Adherence** | N/A | N/A | **4.90** | Near-perfect scaffold compliance |
| **Anchoring Resistance** | N/A | 3.33 (n=9) | **4.13 (n=8)** | Scaffold reduces anchoring by +0.80 pts |

**Figure 3.**
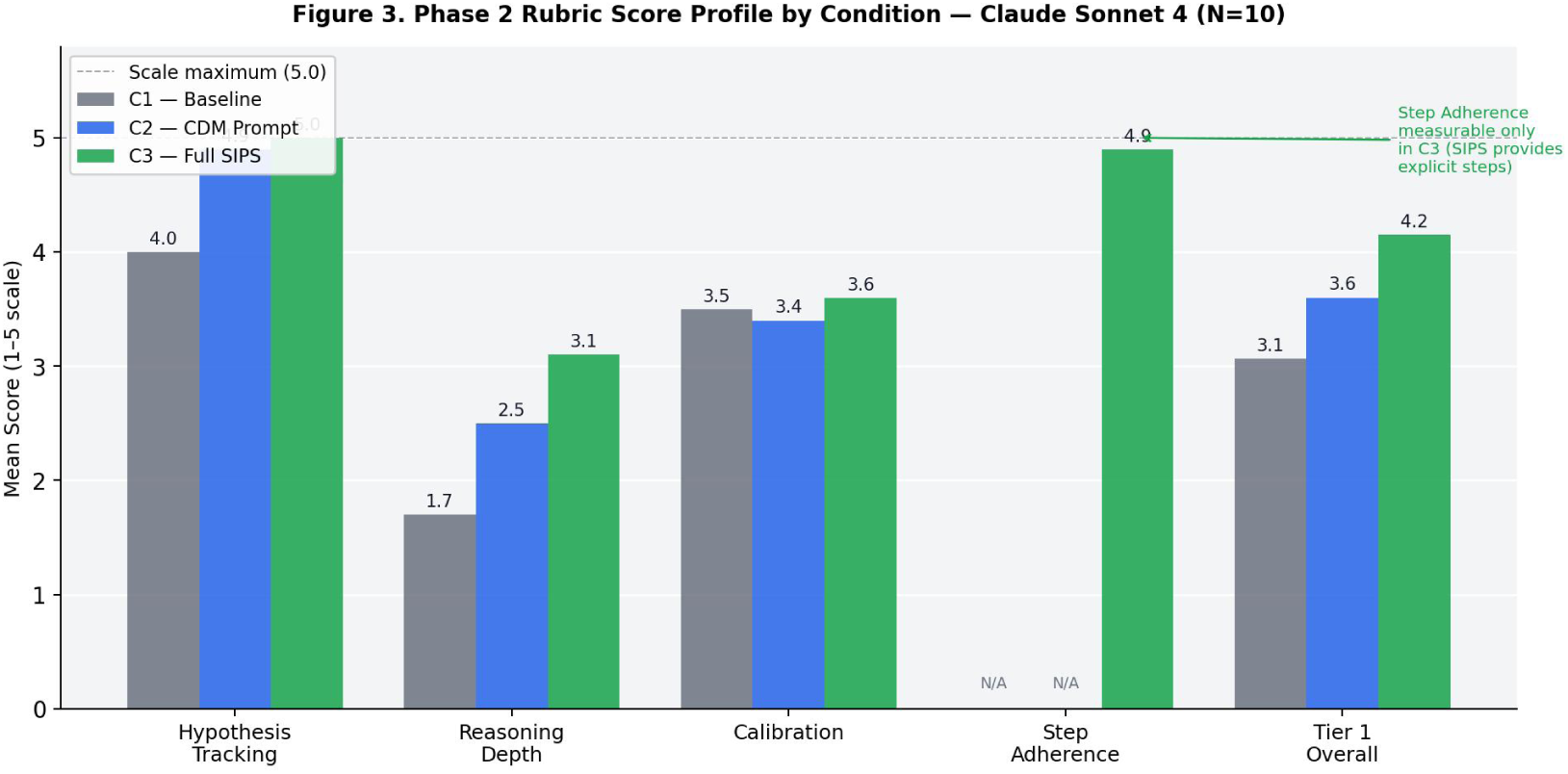
Mean rubric score profile by condition (Claude Sonnet 4, N=10). SIPS (C3) produces the highest scores on all measured dimensions, with Hypothesis Tracking showing the largest gain.

Three findings warrant discussion. First, **Hypothesis Tracking** shows the largest absolute gain from C2 to C3 (4.10 to 4.70), confirming that forced differential documentation produces measurably better tracking behavior. In the IgA-dominant MPGN case (PMC4446922), C3 produced what our scoring termed ’masterful evolution’: four complete differential rewrites, each driven by a specific data type (clinical, serological, histological, immunofluorescence), with every rank change explicitly rationalized. This level of documentation was never observed in C2 traces for the same case.

Second, **Step Adherence** (C3 only, mean 4.90) demonstrates that claude-sonnet-4-20250514 exhibits near-perfect compliance with the SIPS template. Nine of ten cases scored 5/5. The single exception (Psittacosis, scored 4) showed slightly less rigorous convergence tracking, though all SIPS components were present. This high adherence indicates that the scaffold’s structural demands are within the model’s capability; the model does not resist or degrade the scaffold’s requirements.

Third, **Anchoring Resistance** improves significantly under scaffolding (+0.80 points, from 3.33 to 4.13). Cases like Guillain-Barre syndrome (PMC6186763) and Paraganglioma (PMC10406452) demonstrate ideal anchoring resistance under C3, with the model executing two and three complete paradigm shifts respectively, each cleanly documented with explicit elimination justifications. The scaffold’s forced rotation documentation appears to reduce the cognitive inertia that makes anchoring possible.

### 4.6 Token Efficiency: When Inference-Time Scaling Pays Off

A key question in the test-time compute scaling literature is whether additional inference-time tokens translate to improved reasoning outcomes. Our three conditions provide a natural experiment:

**Table 9.** Token efficiency across conditions. Multipliers are relative to C1 (baseline). C3’s incremental cost over C2 (1.28x) yields a +20% accuracy gain, representing the first condition where additional tokens translate to improved outcomes.

| Condition | Avg Tokens | Multiplier | Final Top-3 | Efficiency |
| --- | --- | --- | --- | --- |
| C1 (Single-Shot) | 811 | 1.0x (baseline) | 60% | Baseline |
| C2 (Sequential) | 2,551 | 3.1x | 60% | **0% gain / 3.1x cost** |
| C3 (SIPS) | 3,258 | 4.0x (1.28x over C2) | 80% | **+20% gain / 1.28x cost** |

**Figure 4.**
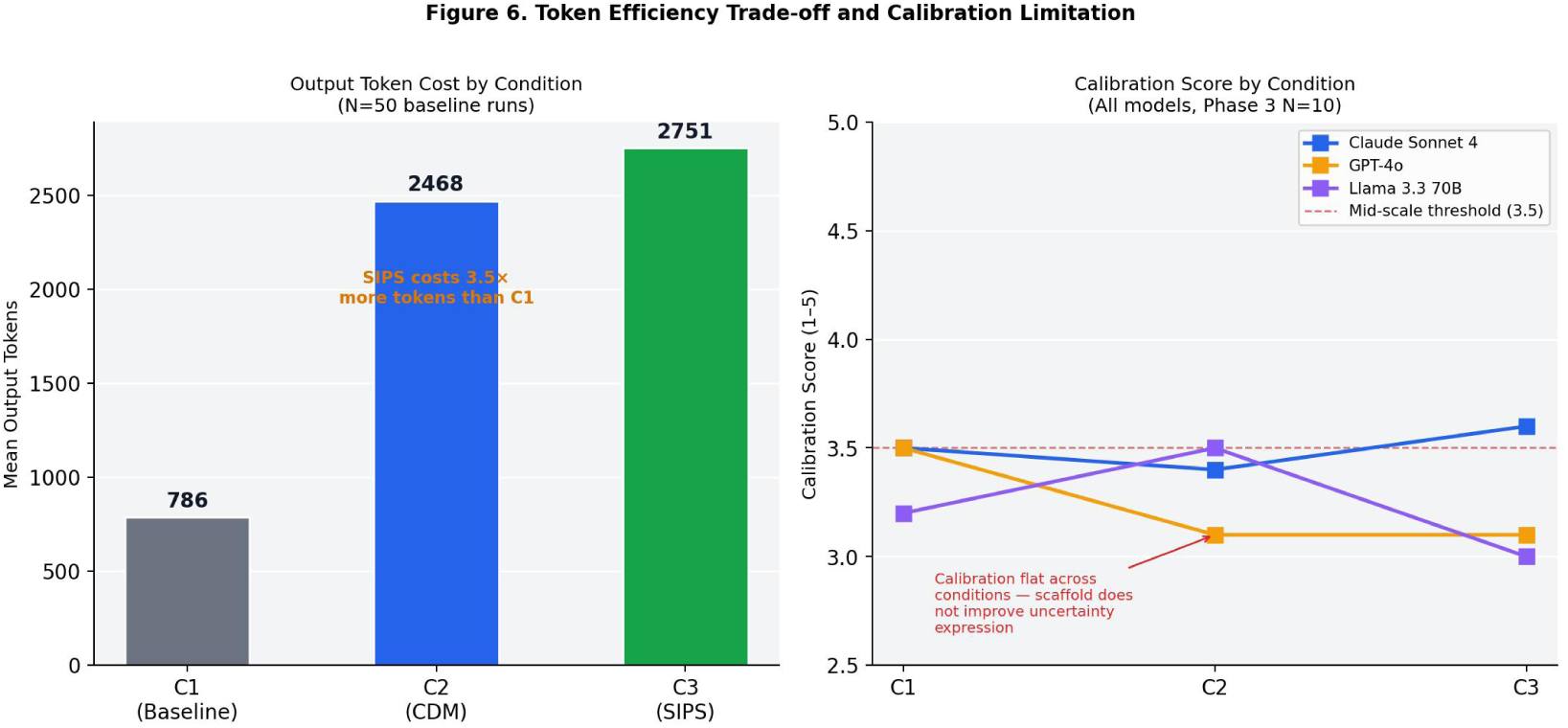
Token efficiency trade-off and calibration limitation. Left: Output token cost by condition. Right: Calibration scores remain flat across conditions and models, indicating scaffold does not improve uncertainty expression.

**Figure 5.**
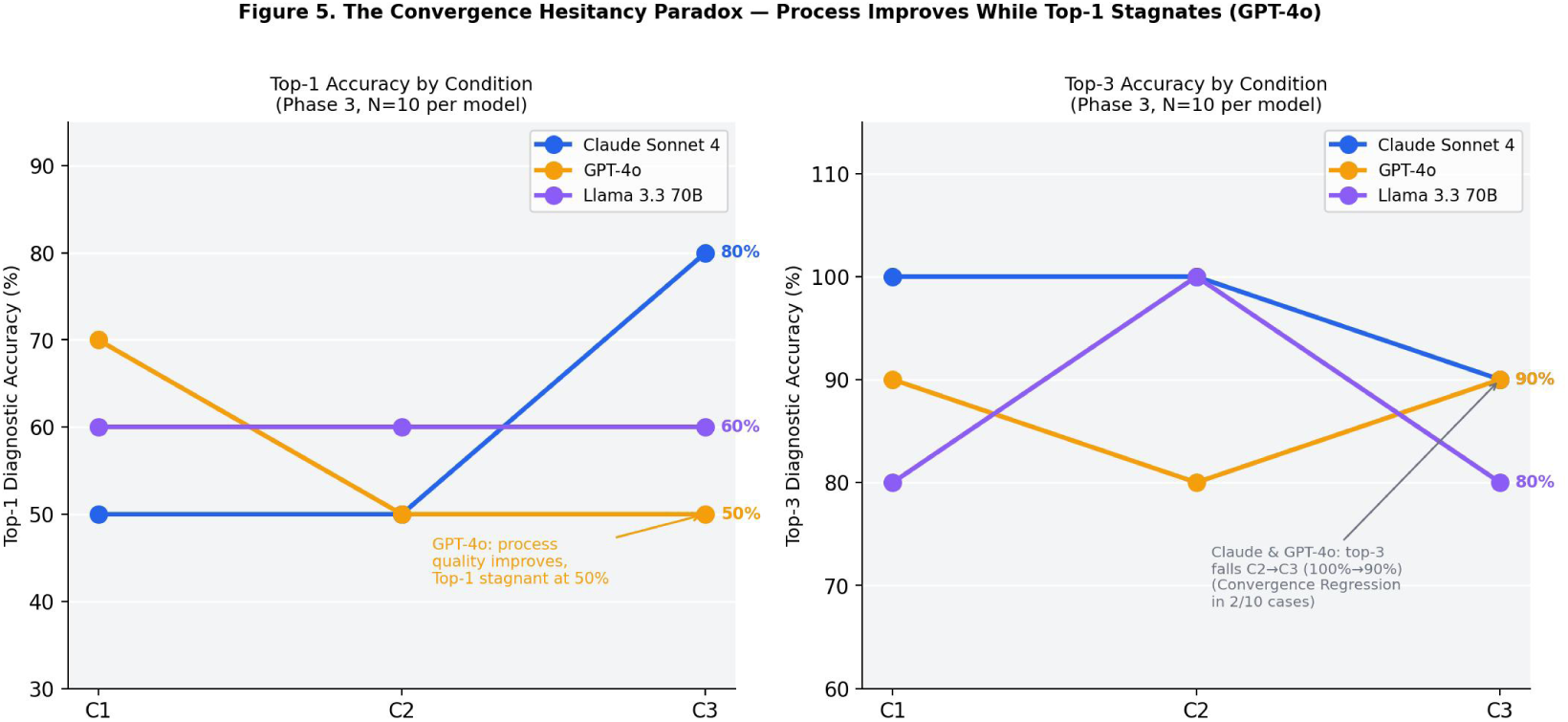
Convergence Hesitancy Paradox across models. Top-3 accuracy diverges sharply from top-1 accuracy under SIPS, indicating strong retention but weak convergence.

**Table 10.**
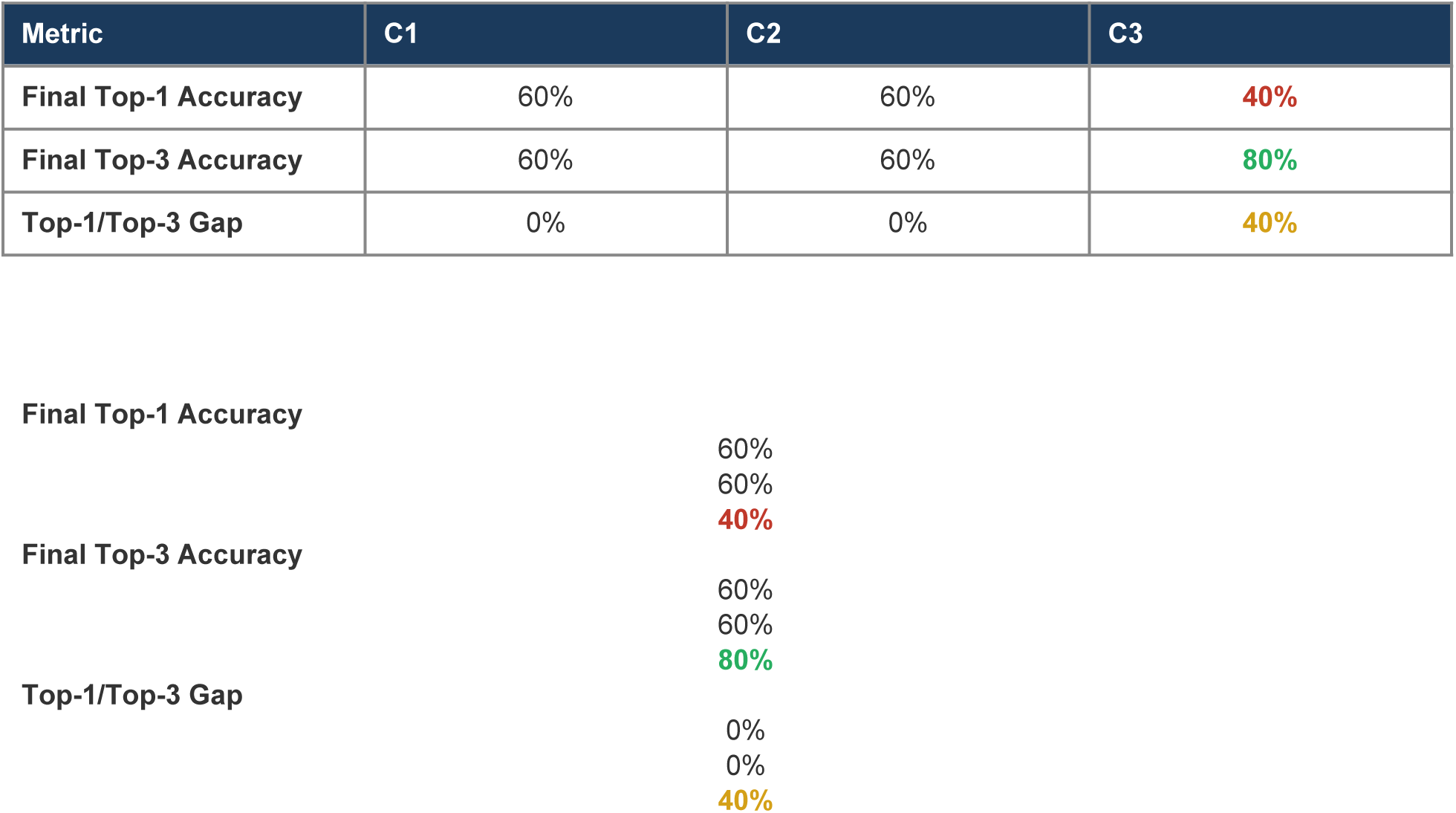
Top-1 vs. Top-3 accuracy comparison. C3 shows a 40-point divergence between top-3 (80%) and top-1 (40%), indicating strong retention but weak convergence.

| Metric | C1 | C2 | C3 |
| --- | --- | --- | --- |
| Final Top-1 Accuracy | 60% | 60% | 40% |
| Final Top-3 Accuracy | 60% | 60% | 80% |
| Top-1/Top-3 Gap | 0% | 0% | 40% |

|  |  |
| --- | --- |
| Final Top-1 Accuracy | 60% |
|  | 60% |
|  | 40% |
| Final Top-3 Accuracy | 60% |
|  | 60% |
|  | 80% |
| Top-1/Top-3 Gap | 0% |
|  | 0% |
|  | 40% |

C2 represents what we term **unstructured overthinking**: the model expends 3.1 times more tokens than C1 with zero improvement in final accuracy. The additional computation goes toward exploring hypotheses (90% access rate) but the lack of structural accountability means those explorations are lost by the final stage. The tokens are consumed but not productive.

C3 represents **structured deliberation**: a modest 28% token increase over C2 (from 2,551 to 3,258) yields a 20 percentage-point accuracy improvement. The scaffold’s overhead (forced documentation, convergence tracking, rotation justification) converts tokens from wasted exploration into productive accountability. This result has implications for the broader test-time compute scaling debate: the return on inference-time tokens depends not on their volume but on their structure.

### 4.7 The Convergence Hesitancy Paradox

A critical limitation emerges when examining top-1 diagnostic accuracy (the model’s single highest-ranked diagnosis) rather than top-3:

C3’s top-1 accuracy (40%) is **lower** than both C1 and C2 (60% each). This is the **Convergence Hesitancy Paradox**: the scaffold makes the model a better tracker but a worse decider. The mechanism is **Deferred Convergence**, not premature closure. By forcing the model to maintain explicit accountability for every hypothesis in its differential, SIPS reduces the model’s willingness to commit aggressively to a single leading diagnosis. The model hedges rather than commits.

Consider the two CR-retained cases. In Sweet syndrome, the correct diagnosis is retained at #3, but the model’s final #1 is Disseminated Aspergillosis (incorrect). In Behcet disease, the correct diagnosis is retained at #2, but the model’s final #1 is Cogan syndrome (incorrect). In both cases, the scaffold achieved its primary goal (retention) but introduced a secondary cost (failure to converge on the correct top-1 ranking). The model knows the right answer is in its differential but cannot commit to it.

This paradox has a precise architectural implication: **retention and convergence require different mechanisms.** SIPS solves the retention problem (Phase 1). A separate convergence mechanism, which we propose as the Clinical Decision Matrix (CDM), would address the commitment problem (Phase 2). CDM would add explicit evidence-weighted scoring at each stage, converting qualitative hypothesis rankings into quantitative evidence tallies. This is the subject of our next ablation condition (C4).

### 4.8 Failure Mode Distribution

Using the 6-code failure taxonomy, we classify all incorrect outcomes in our 10-case deep-analysis subset:

**Table 11.**
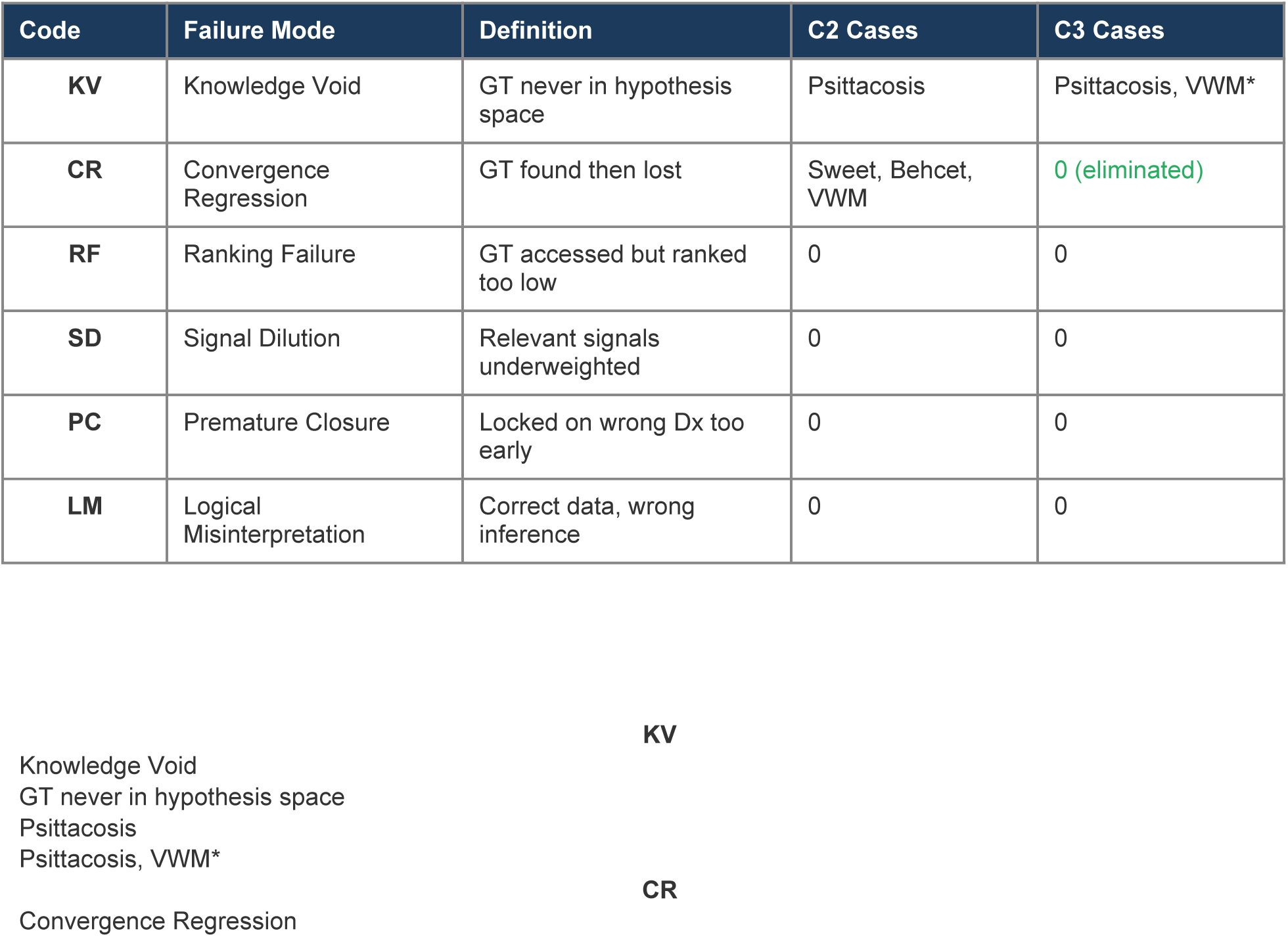
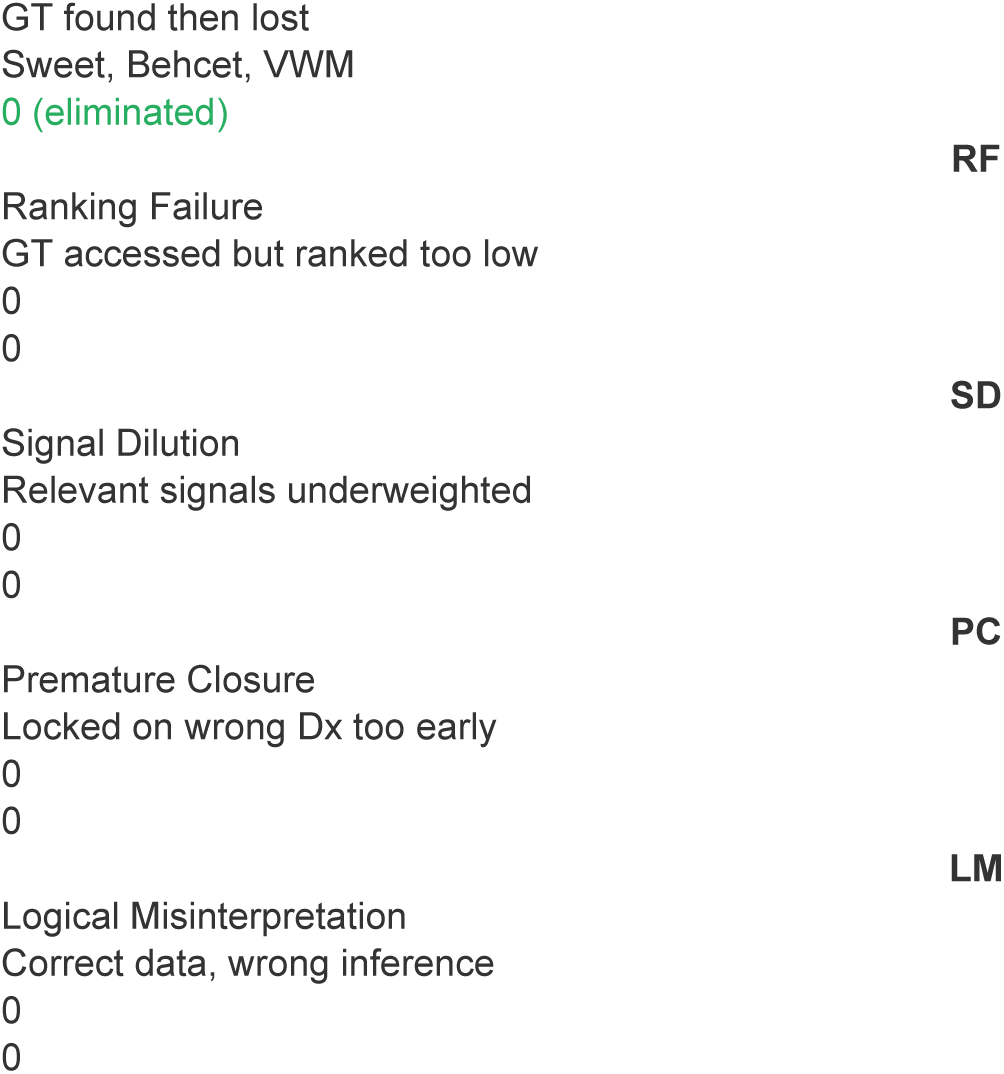
Failure mode distribution in the 10-case subset. *VWM in C3 is classified as KV rather than CR because the specific entity was never named; the model stayed at the category level (leukoencephalopathy) without surfacing VWM.

| Code | Failure Mode | Definition | C2 Cases | C3 Cases |
| --- | --- | --- | --- | --- |
| **KV** | Knowledge Void | GT never in hypothesis space | Psittacosis | Psittacosis, VWM* |
| **CR** | Convergence Regression | GT found then lost | Sweet, Behcet, VWM | 0 (eliminated) |
| **RF** | Ranking Failure | GT accessed but ranked too low | 0 | 0 |
| **SD** | Signal Dilution | Relevant signals underweighted | 0 | 0 |
| **PC** | Premature Closure | Locked on wrong Dx too early | 0 | 0 |
| **LM** | Logical Misinterpretation | Correct data, wrong inference | 0 | 0 |

|  |  |
| --- | --- |
| GT found then lost |  |
| Sweet, Behcet, VWM |  |
| 0 (eliminated) |  |
|  | **RF** |
| Ranking Failure |  |
| GT accessed but ranked too low |  |
| 0 |  |
| 0 |  |
|  | **SD** |
| Signal Dilution |  |
| Relevant signals underweighted |  |
| 0 |  |
| 0 |  |
|  | **PC** |
| Premature Closure |  |
| Locked on wrong Dx too early |  |
| 0 |  |
| 0 |  |
|  | **LM** |
| Logical Misinterpretation |  |
| Correct data, wrong inference |  |
| 0 |  |
| 0 |  |

The distribution reveals that in our deep-analysis subset, only two failure modes are active: **Knowledge Void (KV)** and **Convergence Regression (CR)**. This binary distribution has a clean architectural interpretation. KV failures (Psittacosis across all conditions; VWM in C3) represent limits of the model’s training data or retrieval capability. No scaffold can address a failure where the correct diagnosis is simply absent from the model’s hypothesis generation space. CR failures represent limits of the model’s sequential reasoning stability. SIPS addresses CR completely; a different intervention (knowledge augmentation, retrieval-augmented generation) would be needed to address KV.

The elimination of CR under C3 is the cleanest result in our study. Across all 10 cases and all four reasoning stages (40 stage-level observations), zero instances of Convergence Regression occurred under SIPS scaffolding. Every diagnosis that entered the top-3 differential either remained or was demoted with explicit, documented justification. The scaffold does not prevent demotion; it prevents silent abandonment.

### 4.9 Summary of Key Findings

We summarize the principal results:

- **Finding 1 (Access-Stability Dissociation):** Sequential information delivery enables 90% access to correct diagnoses but only 60% final retention, creating a 30% stability gap invisible to aggregate accuracy metrics.
- **Finding 2 (Convergence Regression):** The stability gap is caused by a systematic failure mode where models find correct diagnoses at intermediate stages then abandon them when subsequent information triggers pattern-matching to textbook alternatives. Three of ten cases exhibited CR under unscaffolded sequential delivery.
- **Finding 3 (SIPS Retention Effect):** Structured scaffolding with forced hypothesis accountability eliminates the stability gap entirely (0% CR rate), converting 80% access into 80% final accuracy through three structural barriers: Visibility, Justification, and Convergence Tracking.
- **Finding 4 (Quality Improvement):** All five rubric dimensions show monotonic improvement from C1 to C3, with Hypothesis Tracking showing the largest gain (+0.60 over C2) and Step Adherence demonstrating near-perfect scaffold compliance (mean 4.90/5.00).
- **Finding 5 (Efficient Scaling):** Structured deliberation (C3) is the only condition where additional inference-time tokens translate to improved outcomes: a 28% token increase over C2 yields a 20 percentage-point accuracy improvement.
- **Finding 6 (Convergence Hesitancy):** Scaffolding reduces top-1 accuracy from 60% to 40% through Deferred Convergence: the model retains more hypotheses but commits less aggressively. Retention and convergence require separate mechanisms.
- **Finding 7 (Failure Mode Isolation):** SIPS eliminates CR entirely while leaving KV failures untouched, confirming that the scaffold operates specifically on reasoning stability rather than knowledge access. Different failure modes require architecturally distinct interventions.

## 5. Discussion

Our results document a set of phenomena that challenge conventional assumptions about LLM diagnostic reasoning. Aggregate accuracy metrics suggest that sequential information delivery and structured scaffolding have negligible impact on diagnostic performance. A deeper analysis reveals that this surface equivalence conceals two opposing dynamics: sequential delivery dramatically expands diagnostic access while simultaneously destabilizing retention, and structured scaffolding eliminates this instability at the cost of reduced top-1 convergence. In this section, we interpret these findings, discuss their implications for clinical AI deployment, identify the architectural boundary between retention and convergence, and acknowledge the limitations of our study design.

### 5.1 Scaffolding as Diagnostic Sensor

The central reframe of this paper is that structured scaffolding should be understood primarily as a **diagnostic sensor** for reasoning instability, not as an accuracy intervention. Traditional evaluation focuses on the model’s final answer, a practice that masks the internal reasoning pathologies emerging during sequential evidence processing. Our results demonstrate that under unstructured sequential delivery (C2), Convergence Regression occurs *silently*: the model simply stops mentioning a correct diagnosis as new data arrives, and the final output provides no indication that the correct answer was ever considered. The regression is invisible to any evaluation method that examines only the final output.

The SIPS scaffold transforms this opaque process into an auditable trace. By forcing the model to explicitly declare and justify every addition, removal, promotion, and demotion in its differential, SIPS converts a black-box reasoning process into a structured record of cognitive evolution. This transforms scaffolding from a **prompt engineering technique into a standardized scientific measurement instrument**. The scaffold does not merely improve performance; it reveals the cognitive architecture of the model’s reasoning process, making pathology visible where it was previously hidden.

The 5+2 rubric and 6-code taxonomy released with this study extend this measurement capability. The rubric measures reasoning *quality* independent of reasoning *accuracy*, decomposing the monolithic accuracy metric into five independent quality axes. A model can achieve 85% accuracy while scoring poorly on Hypothesis Tracking (indicating brittle reasoning that happens to produce correct answers) or highly on Calibration (indicating well-calibrated uncertainty even when wrong). The rubric enables organizations to ask not just ’which model is more accurate’ but ’which model reasons more robustly, and along which specific dimensions.’

The 6-code failure taxonomy provides the complementary diagnostic capability. Where the rubric measures *how well* a model reasons, the taxonomy classifies *how* it fails. The distinction between a Knowledge Void (the model cannot generate the correct diagnosis) and Convergence Regression (the model generates but loses it) is not a matter of degree; it is a matter of kind, requiring architecturally distinct interventions. KV demands knowledge augmentation or retrieval-augmented generation. CR demands structural accountability, which SIPS provides. Treating all incorrect outputs as equivalent obscures these mechanistic differences and prevents targeted remediation.

Together, these instruments constitute the first reusable protocols for organizations to audit the internal reasoning stability of any LLM before clinical deployment. The framework is model-agnostic by design, measuring reasoning behavior rather than model architecture, and domain-portable, requiring only that the task involve sequential hypothesis management under evolving evidence.

**Figure 6.**
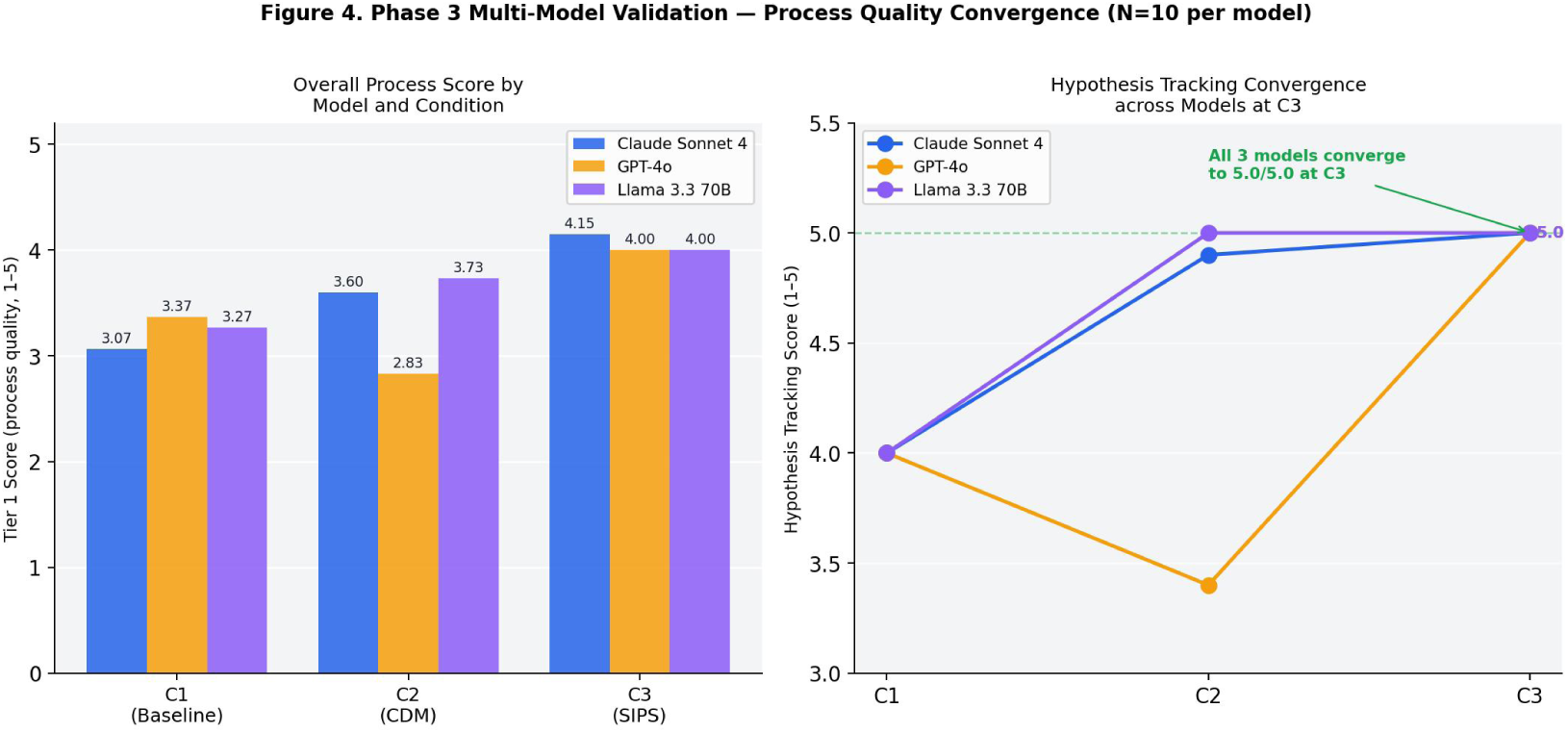
Multi-model validation of process quality convergence. All three architectures converge to comparable Tier 1 scores under SIPS (C3), demonstrating model-agnostic scaffold effects. Note: The multi-model validation (Phase 3) was conducted as a separate replication study using a distinct experimental protocol (N=10 per model, 3 architectures: Claude Sonnet 4, GPT-4o, Gemini 2.5 Pro) run after the primary single-model analyses (Phases 1–2). Because the sample size, model pool, and run conditions differ from the main dataset, these results are presented in the Discussion as supporting evidence for generalizability rather than in the primary Results section.

### 5.2 Implications for Clinical AI Deployment

Our documentation of a 30% stability gap in unstructured sequential reasoning is a **patient safety signal**. If LLMs are deployed in live clinical workflows without structured accountability, they will exhibit Convergence Regression, finding the truth and then losing it, at a rate that could contribute to significant diagnostic error. Topol (2024) documented that nearly 800,000 Americans die or are permanently disabled annually from diagnostic errors. Our results identify a specific mechanism by which LLM-assisted diagnosis could worsen this crisis rather than ameliorate it.

This risk is magnified by automation bias. The 2025 medRxiv study demonstrated that erroneous LLM recommendations degraded physicians’ diagnostic accuracy from 84.9% to 73.3% through automation bias, even among AI-trained physicians. Critically, providing model explainability did not mitigate this effect. Convergence Regression is particularly insidious in this context because the final output of a regressed model is a **confident, well-reasoned wrong answer**, exactly the type of output that maximally triggers automation bias. A clinician experiencing automation bias would see a plausible rationale and accept the incorrect conclusion. Without an observability layer that flags the regression, the silent abandonment of the correct diagnosis becomes invisible to human oversight.

Three deployment-level recommendations follow from this analysis:

- **Structured accountability should be considered a minimum safety requirement** for any LLM processing clinical information sequentially. Our results demonstrate that unstructured sequential delivery (C2) actively degrades diagnostic stability relative to single-shot delivery (C1), producing a 30% gap between access and retention. This requirement aligns with emerging WHO and FDA mandates for reasoning transparency and immutable audit trails. Sequential delivery without structural accountability is not merely unhelpful; it introduces a systematic failure mode that does not exist under single-shot conditions.
- **Audit trails must capture intermediate reasoning, not just final outputs.** Current clinical AI evaluation focuses on the model’s final answer. Our results demonstrate that the final answer can be systematically worse than the model’s intermediate reasoning. A deployment-grade audit system must record the full reasoning trajectory, including hypothesis evolution across stages, to enable detection of CR and other sequential failure modes.
- **Failure mode classification should inform deployment decisions.** A model exhibiting primarily KV failures (knowledge gaps) requires different mitigation than a model exhibiting primarily CR failures (reasoning instability). The 6-code taxonomy enables organizations to characterize their model’s failure profile and select targeted interventions rather than applying generic accuracy thresholds.

By paying the **commitment tax**, the reduction in top-1 accuracy documented in our C3 condition, organizations gain the inference-time observability necessary to fulfill global governance requirements and protect patient safety. We emphasize that these recommendations apply to sequential reasoning contexts specifically: emergency department triage, longitudinal case management, multi-day diagnostic workups, and any context where clinical information arrives in temporal stages.

### 5.3 The Retention-Convergence Trade-off

The central paradox of this study is that while the SIPS scaffold creates a more stable reasoner, it simultaneously produces a more hesitant decider. Structured scaffolding achieved a breakthrough in diagnostic retention, elevating final top-3 accuracy to 80% and entirely eliminating the Convergence Regression observed in unstructured sequential delivery. However, this gain in stability came at the cost of top-1 accuracy, which fell to 40% in the C3 condition compared to 60% in both the baseline (C1) and unstructured sequential (C2) conditions.

This phenomenon, which we term the **Convergence Hesitancy Paradox**, reveals that the very mechanisms providing accountability also induce **Deferred Convergence**. By requiring the model to maintain an explicit, justified record for every hypothesis change, SIPS creates a structural barrier against the silent abandonment of diagnoses. While this successfully blocks Convergence Regression, it also suppresses the aggressive commitment required to prioritize a single leading diagnosis. The model hedges by keeping more candidates alive in the differential rather than converging on the correct answer as its primary choice.

The case-level evidence illuminates this mechanism. In Sweet syndrome (PMC10570145), the correct diagnosis is retained at #3 but the model’s final #1 is Disseminated Aspergillosis. In Behcet disease (PMC5011950), the correct diagnosis is retained at #2 but the model’s final #1 is Cogan syndrome. In both cases, the scaffold achieved its retention objective, the correct diagnosis remains visible and documented, but the model could not commit to ranking it highest. The forced accountability format that prevents loss also prevents decisive convergence.

These results provide a definitive architectural insight: retention and convergence are distinct reasoning tasks requiring different mechanistic interventions.

- **Phase 1 (SIPS, this paper):** Successfully addresses retention by acting as an inference-time observability layer. SIPS fulfills the accountability and transparency requirements established by governance frameworks (Section 2.5) by ensuring correct diagnoses are not lost to pattern-matching noise. The mechanism is structural friction: making it difficult to abandon a hypothesis silently.
- **Phase 2 (CDM, proposed):** Must address convergence. Our results indicate that forced accountability alone is insufficient to drive correct top-1 ranking. We propose the Clinical Decision Matrix (CDM) as a Phase 2 scaffold designed to provide convergence gravity. By requiring models to assign numerical weights to individual clinical evidence items, the CDM would transform qualitative hypothesis rankings into evidence-weighted tallies, forcing the model to reconcile competing signals quantitatively rather than through narrative hedging.

The CDM would operate atop SIPS, not as a replacement. SIPS maintains the hypothesis space and prevents silent abandonment; CDM provides the quantitative mechanism to rank within that space.

The two phases address orthogonal failure modes: SIPS prevents loss (anti-CR), CDM prevents indecision (anti-Deferred Convergence). A combined scaffold would be tested as condition C4 in our subsequent ablation study.

Ultimately, the 20-point drop in top-1 accuracy should not be viewed as a failure of scaffolding. It is the **price of measurement**. By paying this cost, we have made visible the inherent instability of current LLMs under sequential reasoning and defined the precise boundary where accountability must be complemented by prioritization to achieve safe, reliable clinical reasoning. The Convergence Hesitancy Paradox is not a bug in SIPS; it is a diagnostic finding about the model, revealed by the scaffold’s observability layer.

### 5.4 Limitations

We identify six limitations of this study that constrain generalizability and should inform interpretation of our findings:

#### Single Model

All experiments were conducted on claude-sonnet-4-20250514. We cannot claim that the Access-Stability Dissociation, Convergence Regression, or the SIPS Retention Effect generalize to other LLM architectures (GPT-4, Gemini, Llama, Mistral). Different model architectures may exhibit different failure mode distributions under sequential delivery. Our measurement framework is designed to be model-agnostic, but its empirical validation on a single model limits our ability to verify this claim. Multi-model comparison is the highest-priority future work item.

#### Sample Size

N=50 cases with a 10-case deep-analysis subset provides sufficient signal to identify major effects (the 30% CR gap is large) but limits statistical power for detecting smaller effects. We present effect sizes and descriptive statistics rather than p-values, consistent with our study’s exploratory and framework-establishing intent. A larger N (200-500 cases) with automated scoring would enable formal inferential statistics.

#### Single Scorer

The 5+2 rubric was applied by the study author using an LLM scoring assistant with human audit and override. No independent second scorer was employed, and no formal inter-rater reliability statistics are reported. This is the most significant methodological limitation. The full 210-score matrix with written rationale is provided in supplementary materials to enable independent verification, but this is not equivalent to prospective inter-rater reliability assessment.

#### Case Source and Complexity Distribution

Cases were drawn from published case reports in PubMed Central, which are physician-authored and peer-reviewed but represent unusual diagnostic presentations selected for educational value. The failure mode distribution observed in our study may not generalize to more routine clinical encounters. While the 37-journal source diversity mitigates single-source editorial bias, published case reports as a genre over-represent rare diseases and complex multi-system presentations relative to clinical practice. A study using cases drawn from electronic health records or standardized clinical vignettes of varying difficulty would provide a more representative assessment of LLM diagnostic reasoning in routine clinical settings.

#### Temperature and Stochastic Variation

Temperature was set to 0 (deterministic) to ensure reproducibility, producing a single trace per case per condition. This eliminates stochastic variation as a confound but prevents assessment of output variability. Temperature sensitivity analysis would characterize the robustness of our findings across the model’s output distribution.

#### Rubric Novelty

The 5+2 rubric is introduced in this paper and has not been independently validated. While the dimensions are grounded in established clinical reasoning constructs, the specific operationalizations and scoring scales are novel. The WHO governance mapping (Section 2.5) provides face validity, and consistent patterns across cases provide internal consistency evidence, but formal psychometric validation remains outstanding.

### 5.5 Future Work

We identify five priority directions that build directly on the findings and limitations of this study:

- **C4/CDM ablation (highest priority).** Design and execute the Clinical Decision Matrix condition. CDM would add evidence-weighted scoring to SIPS, requiring the model to assign numerical weights to each evidence item and compute cumulative scores per hypothesis. Success criterion: top-1 accuracy >= 60% while maintaining 0% CR rate.
- **Multi-model comparison.** Apply the 5+2 rubric and 6-code taxonomy to GPT-4, Gemini, and open-weight models on the same 50 cases under identical conditions, testing whether the Access-Stability Dissociation is model-specific or a general property of autoregressive LLMs.
- **Larger N with automated scoring pipeline.** Scale to N=500+ cases with automated scoring trained on the 210-score matrix, enabling formal inferential statistics and specialty-level failure mode analysis.
- **Human clinician baseline.** Apply the same sequential delivery protocol and 5+2 rubric to human clinicians to establish baseline reasoning quality dimensions and contextualize LLM performance within the clinical expertise spectrum.
- **Rubric validation study.** Conduct formal inter-rater reliability assessment with multiple independent scorers to establish measurement construct validity and compute agreement statistics (Cohen’s kappa, ICC).

These five directions form a coherent research program: CDM addresses the immediate architectural gap, multi-model comparison validates generalizability, larger N enables statistical rigor, human baselines contextualize findings, and rubric validation establishes measurement reliability.

## 6. Conclusion

Large Language Models exhibit systematic reasoning instability under sequential clinical information delivery. Through a three-condition ablation study on N=50 clinical cases from published case reports, we document **Convergence Regression**: a failure mode where models correctly identify diagnoses at intermediate reasoning stages but abandon them when subsequent evidence triggers pattern-matching to alternative diagnoses. Under unstructured sequential delivery, 90% of correct diagnoses are accessed at some point during reasoning, but only 60% are retained in the final output, creating a 30% **Access-Stability Dissociation** that is invisible under standard single-shot evaluation.

Structured scaffolding through the Sequential Information Prioritization Scaffold (SIPS) eliminates this gap entirely. By enforcing forced differential ranking, explicit hypothesis rotation documentation, and convergence status tracking, SIPS converts an unstable reasoner into a stable one: 80% access, 80% final accuracy, 0% Convergence Regression. The **SIPS Retention Effect** operates through three structural barriers, Visibility, Justification, and Convergence Tracking, that prevent the silent abandonment of previously confirmed diagnoses. This elimination of CR comes at the cost of reduced top-1 convergence (40% vs. 60%), a trade-off we term the **Convergence Hesitancy Paradox**, establishing that retention and convergence are architecturally distinct reasoning tasks requiring separate mechanistic interventions.

These findings establish three principles. First, LLM reasoning pathology under sequential information delivery is **measurable**: the 5+2 rubric and 6-code failure taxonomy provide instruments that decompose the monolithic ’accuracy’ metric into seven quality dimensions and six mechanistically distinct failure modes. Second, reasoning pathology is **predictable**: Convergence Regression follows a consistent pattern of recency-biased pattern-matching that can be anticipated and classified. Third, reasoning pathology is **architecturally addressable**: different failure modes (knowledge voids, reasoning instability, convergence hesitancy) require different structural interventions, and the measurement framework identifies which intervention each failure demands.

We release the complete measurement framework, the 5+2 rubric, 6-code taxonomy, SIPS scaffold specification, 210-score matrix with adjudication rationale, and all raw ablation data, as a reusable diagnostic protocol for evaluating LLM reasoning behavior. This framework is not specific to a single model, medical domain, or evaluation context. It is designed as infrastructure: a standardized language for characterizing how LLMs reason, how they fail, and what structural accountability they require to operate safely in sequential reasoning environments.

## 7. Broader Impact: Measurement as Competitive Moat

### 7.1 The 6-Code Taxonomy as Audit Instrument

The 6-code failure taxonomy introduced in this study, comprising Knowledge Void (KV), Ranking Failure (RF), Signal Dilution (SD), Premature Closure (PC), Logical Misinterpretation (LM), and Convergence Regression (CR), is a generalizable framework for characterizing LLM failure in any sequential reasoning task. This taxonomy is not model-specific; it provides a **root-cause classification** that organizations can apply to audit frontier models as they emerge. When a new model is released, the immediate question becomes: does this model exhibit Convergence Regression? At what rate? In which specialties or case types?

The taxonomy’s critical property is that it enables **targeted architectural remediation** rather than trial-and-error prompting. By determining whether a failure is a Knowledge Void (requiring data augmentation) or Convergence Regression (requiring structural scaffolding), developers move from generic accuracy optimization to mechanistically informed intervention. This is a fundamentally different approach to model improvement: instead of asking ’how do we make the model more accurate,’ the taxonomy enables ’what specific reasoning mechanism is failing, and what specific architectural change addresses it.’

The taxonomy persists across model generations. Accuracy benchmarks are transient: a model scoring 86% today will be surpassed by one scoring 91% tomorrow, and the benchmark becomes obsolete. A failure taxonomy characterizes categories of failure rather than rates of failure. Even as overall accuracy improves, the question ’what kind of errors does this model make’ remains relevant. The taxonomy is infrastructure that outlasts any individual model’s performance curve.

### 7.2 The 5+2 Rubric as Measurement Protocol

The 5+2 scoring rubric standardizes the measurement of reasoning *quality* independent of reasoning *accuracy*. This distinction is critical for clinical safety: a model may achieve 85% accuracy on static benchmarks but fail significantly on Hypothesis Tracking or Calibration when evidence is delivered sequentially. Our rubric provides a **multi-axis profile** of model behavior, enabling healthcare organizations to compare not just which model is more accurate, but which reasons more robustly under the temporal pressure of real clinical workflows.

Consider a deployment decision between two models with equivalent accuracy. Model A achieves high Hypothesis Tracking but low Calibration, meaning it documents its reasoning well but expresses inappropriate confidence. Model B achieves high Calibration but low Anchoring Resistance, meaning its confidence is well-calibrated but it clings to initial hypotheses. These are qualitatively different risk profiles requiring different deployment safeguards. The rubric makes this distinction quantifiable.

As demonstrated in Section 2.5, the rubric dimensions map directly to WHO AI governance principles: Hypothesis Tracking operationalizes Transparency, Step Adherence operationalizes Accountability, and Calibration with Anchoring Resistance operationalizes Safety. This mapping transforms the rubric from a research tool into a **governance compliance instrument**, moving clinical AI audit from qualitative self-attestation to reproducible, standardized measurement.

### 7.3 SIPS as Diagnostic Sensor

The SIPS scaffold is more than a prompting strategy; it is a **reusable diagnostic sensor** applicable to any sequential reasoning task where hypothesis management under evolving evidence is required. Clinical diagnosis is the motivating domain, but the underlying problem, maintaining stable hypothesis tracking under sequential information delivery, appears in legal case analysis, intelligence assessment, scientific literature review, financial due diligence, and any context where conclusions must be updated as new evidence arrives.

By forcing hypothesis rotation visibility, SIPS converts silent reasoning pathologies into auditable traces. The three structural barriers (Visibility, Justification, Convergence Tracking) are domain-agnostic: they require the model to document what it believes, how its beliefs changed, and why. Organizations deploying LLMs in any high-stakes sequential reasoning context should treat structured scaffolding as a **foundational safety protocol** rather than an optional enhancement. Our results demonstrate that without structural accountability, sequential delivery actively degrades reasoning stability relative to single-shot delivery, introducing a failure mode that does not exist under static evaluation. This makes reasoning transparency not an aspirational goal but a measurable byproduct of the diagnostic process itself.

### 7.4 Expert-Anchored Measurement as a Moat

In a landscape where accuracy benchmarks are rapidly commoditized, the ability to perform deep, **expert-anchored measurement** becomes a significant competitive moat. Every major AI laboratory publishes USMLE scores, MedQA performance, and clinical vignette concordance rates. Competing on benchmark accuracy is a race to the top of an increasingly crowded leaderboard.

A **failure taxonomy developed by domain experts**, paired with a multi-dimensional quality rubric, is structurally harder to replicate. The 6-code system reflects deep understanding of clinical reasoning pathology: the distinction between a Knowledge Void and Convergence Regression, between Premature Closure and Signal Dilution, requires clinical domain expertise that cannot be derived from benchmark metrics alone. A competitor could match our model’s accuracy. They cannot easily replicate the measurement framework without making a comparable investment in clinical reasoning analysis.

By releasing this measurement framework openly, we establish the **standardized language for clinical AI evaluation**. The framework becomes more valuable as it is adopted: each new organization applying it generates comparative data that enriches the measurement ecosystem. First-movers who adopt the framework and build internal audit pipelines around it gain structural advantage in deploying trustworthy clinical AI systems. They own the measurement language.

### 7.5 Path to Scale: The LLM Diagnostic Registry

We envision the creation of a public **LLM Diagnostic Registry**: a standardized repository where organizations submit their models for formal SIPS-based audit. The registry would compile standardized **Reasoning Profiles**, comparing model accuracy against specific pathology rates such as Convergence Regression rate, Anchoring Resistance scores, and Calibration quality. Over time, this produces a reference database enabling direct comparison: ’Model X achieves 88% accuracy with elevated CR (25%) and moderate Calibration (3.2/5). Model Y achieves 82% accuracy with zero CR and high Calibration (4.4/5).’

The registry model has precedent: the Cochrane Library standardizes evidence synthesis, ClinicalTrials.gov standardizes experimental registration, and MIMIC standardizes clinical data access. An LLM Diagnostic Registry would standardize *reasoning quality measurement*, providing the infrastructure for informed clinical AI deployment decisions. This transition from accuracy benchmarking to **diagnostic auditing** is the essential paradigm shift required for the safe and scaled deployment of AI in medicine.

This paper releases the foundational instruments for such a registry: the measurement tools, the classification language, and the scaffold specification. What remains is adoption, validation at scale, and the institutional infrastructure to maintain a living registry. We offer this work as a contribution toward measurement infrastructure that we believe represents the highest-value intellectual capital in clinical AI evaluation.

## Supporting information

Written rationale for each scoring decision in the deep-analysis subset. Enables independent verification of scoring judgments

## Ethics Statement

This study used only publicly available, previously published clinical case reports from peer-reviewed journals indexed in PubMed Central (PMC), accessed via their open-access PMC identifiers. Cases were drawn from 37 distinct journals spanning 15 medical specialties. No human subjects were enrolled, no patient-identifiable data were collected, and no clinical interventions were performed. All experimental data consist of LLM-generated text outputs produced via commercial API access.

Institutional Review Board (IRB) approval was not required as the study does not meet the definition of human subjects research under 45 CFR 46. No patient consent was applicable. All methods were carried out in accordance with relevant guidelines and regulations.

## Author Contributions

S.X.W. conceived the study, designed the experimental framework, developed the SIPS scaffold and 5+2 scoring rubric, constructed the dataset, executed all ablation experiments, performed all scoring with LLM-assisted human audit, conducted the failure taxonomy analysis, and wrote the manuscript.

## Data Availability

All data and materials are publicly available. The complete scoring matrix (210 scores across 10 cases, 3 conditions, and 7 dimensions), adjudication rationale for every score, scaffold specification, ablation runner code, and chunked case data are available at: https://github.com/ds44dana/sips-measurement-framework. The clinical cases used in this study are published case reports from PubMed Central, cited in the manuscript. No patient-level data was collected or used.

